# The Impact of COVID-19 Pandemic on Mental Health: A Scoping Review

**DOI:** 10.1101/2022.12.12.22283388

**Authors:** Blessing O. Josiah, France Ncube

## Abstract

**Background:** This scoping review assessed the COVID-19 impacts on mental health and associated risk factors.

**Methods:** A systematic literature search for relevant articles published in the period March 2020 to July 2022, was conducted in the APA PsychInfo, JBI Evidence Synthesis, Epistemonikos, PubMed, and Cochrane databases.

**Results:** A total of 72 studies met the inclusion criteria. Results showed that the commonly used mental health assessment tools were the Patient Health Questionnaire (41.7%), Generalized Anxiety Disorder Scale (36%), 21-item Depression, Anxiety, and Stress (13.9%), Impact of Event Scale (12.5%), Pittsburgh Sleep Quality Index (9.7%), Symptom Checklist and the General Health Questionnaire (6.9% each). The prevalence rate of depression ranged from 5-76.5%, 5.6-80.5% for anxiety, 9.1-65% for Post-Traumatic Stress Disorder, 8.3-61.7% for sleep disorders, 4.9-70.1% for stress, 7-71.5% for psychological distress, and 21.4-69.3% for general mental health conditions. The major risks included female gender, healthcare-related/frontline jobs, isolation/quarantine, poverty, lower education, COVID-19 risk, age, commodities, mental illness history, negative psychology, and higher social media exposure. The incidence of mental disorders increased along with the increasing cases of COVID-19 and the corresponding government restrictions.

**Conclusion:** Standard assessment tools were used for mental health assessment by the reviewed studies which were conducted during COVID-19. Mental health disorders like depression, anxiety, and stress increased during the COVID-19 pandemic and lockdowns. Various factors impacted the prevalence of mental health disorders. Policymakers need to provide social protective measures to improve coping capacities during critical health events to avoid negative impacts on the population. Further studies should investigate the effectiveness of interventions for reducing the prevalence and risk factors for mental health conditions during a public health challenge.

**Background:** 

## Introduction

The COVID-19 outbreak was declared a pandemic in 2020, with over half a billion cases and over 6 million deaths by the end of May 2022 (1). The high transmissibility, morbidity, and mortality rate led governments to adopt strict measures such as quarantines, restrictions on social gatherings, travel, and closure of borders, schools, churches, and workplaces which disrupted the lives of many people (2,3). COVID-19-related factors, especially the response measures, induced significant levels of stress among people (4–6). An increase in mental health conditions has been recently observed due to these disruptions and stresses (2,7).

Some review studies have been conducted on the pandemic’s impact on mental health in the general population, including a systematic review by Xiong et al. which used studies conducted before the 18^th^ of May 2020 (8). There is also another study conducted by Hannemann et al., but the study population was limited only to medical staff (9). Another scoping review was conducted on the impact of the pandemic on people with similar mental health conditions (10), however, the review only included evidence and literature from the first year of the pandemic (i.e., 2020). Similarly, a scoping review (11) conducted among children and young people only included evidence from the early stage of the pandemic. Meanwhile, all the studies identified pointed toward an occurrence or expectation of a heightened prevalence of mental disorders during the COVID-19 pandemic.

A study conducted in the United States found that almost half of the study participants experienced high levels of anxiety and stress three months into the COVID-19 pandemic (12). About one-third of the population in the UK likewise experienced high levels of anxiety according to the Office for National Statistics (ONS) (13). In Italy, several people suffered from COVID-19-related stress, severe anxiety, and insomnia (6). Although several primary studies have investigated these mental health conditions, there is a lack of recent scoping reviews to summarise findings on this key concept. Consequently, this review summarises findings on the assessment tools, prevalence, risk factors, and trends of mental health conditions during the COVID-19 pandemic.

## Methods

This scoping review was conducted in accordance with the Preferred Reporting Items for Systematic Review and Meta-Analyses (PRISMA) guidelines (14), and recommendations of the Joanna Briggs Institute Manual for Evidence Synthesis (15), and similar studies for the protocol design (16,17).

### Identification of relevant studies

The APA PsychInfo, JBI Evidence Synthesis, Epistemonikos, PubMed, and Cochrane databases were searched to identify relevant articles (Table 1). A final search string with truncations was (mental health* OR mental illness* OR psychiatric situation* OR sanity* OR psychological* OR psychiatric disorder* OR mental health condition* OR mental health disorder* OR mental disease* OR mental stress*) AND (COVID-19 OR COVID-19 pandemic* OR pandemic* OR COVID-19 outbreak* OR lockdown measures* OR Coronavirus* OR SARS-COV2 OR epidemic*) AND (impact* OR effect* OR cause* OR influence* OR result of OR challenge*). In addition, a manual search was done on the references of the most relevant peer-reviewed papers to gather more results suitable for this study (Fig 1).

**Table 1:**
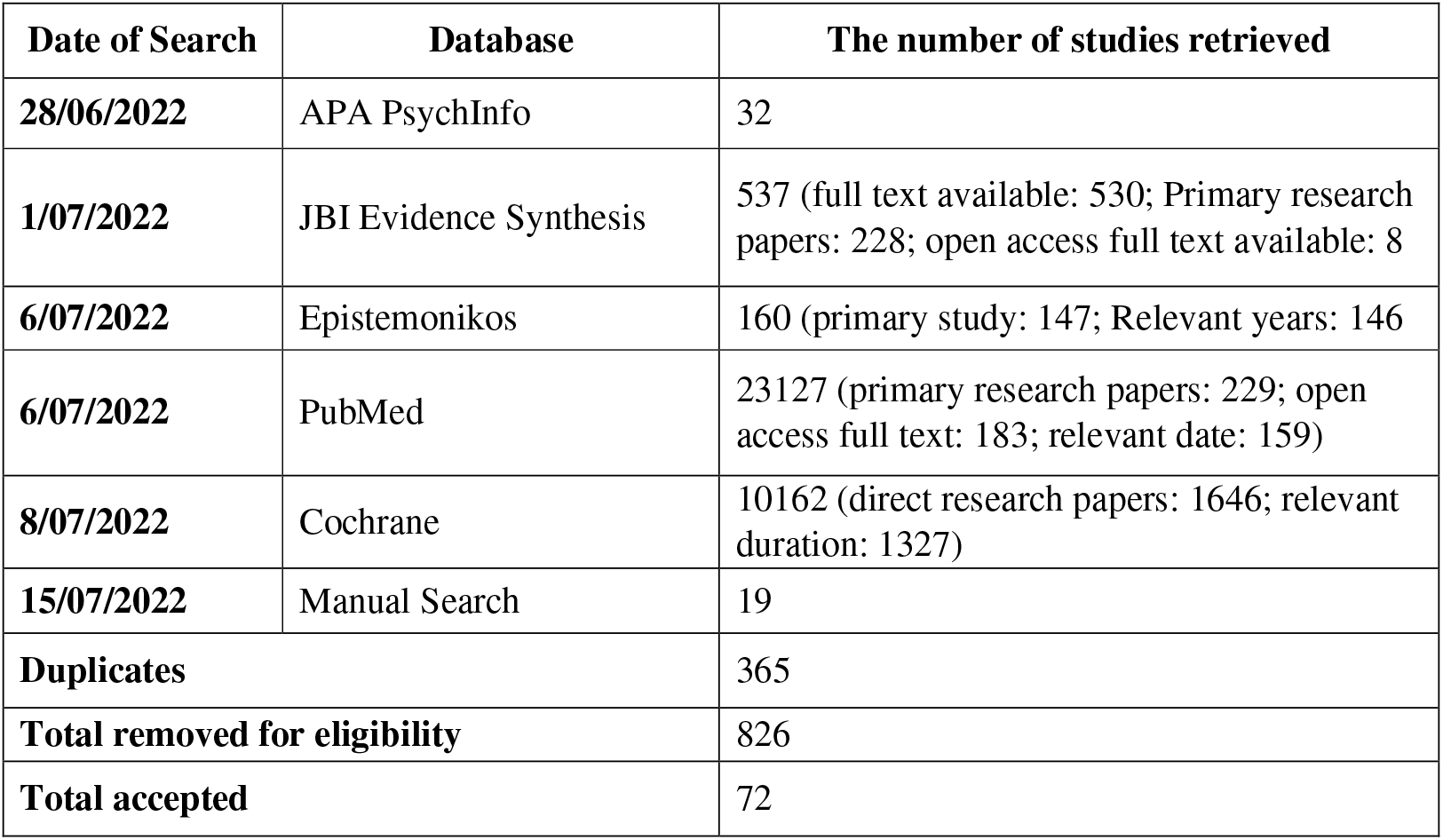
Table of search strategies.

| **Date of Search** | **Database** | **The number of studies retrieved** |
| --- | --- | --- |
| **28/06/2022** | APA PsychInfo | 32 |
| **1/07/2022** | JBI Evidence Synthesis | 537 (full text available: 530; Primary research papers: 228; open access full text available: 8) |
| **6/07/2022** | Epistemonikos | 160 (primary study: 147; Relevant years: 146) |
| **6/07/2022** | PubMed | 23127 (primary research papers: 229; open access full text: 183; relevant date: 159) |
| **8/07/2022** | Cochrane | 10162 (direct research papers: 1646; relevant duration: 1327) |
| **15/07/2022** | Manual Search | 19 |
| **Duplicates** |  | 365 |
| **Total removed for eligibility** |  | 826 |
| **Total accepted** |  | 72 |

**Fig 1:**
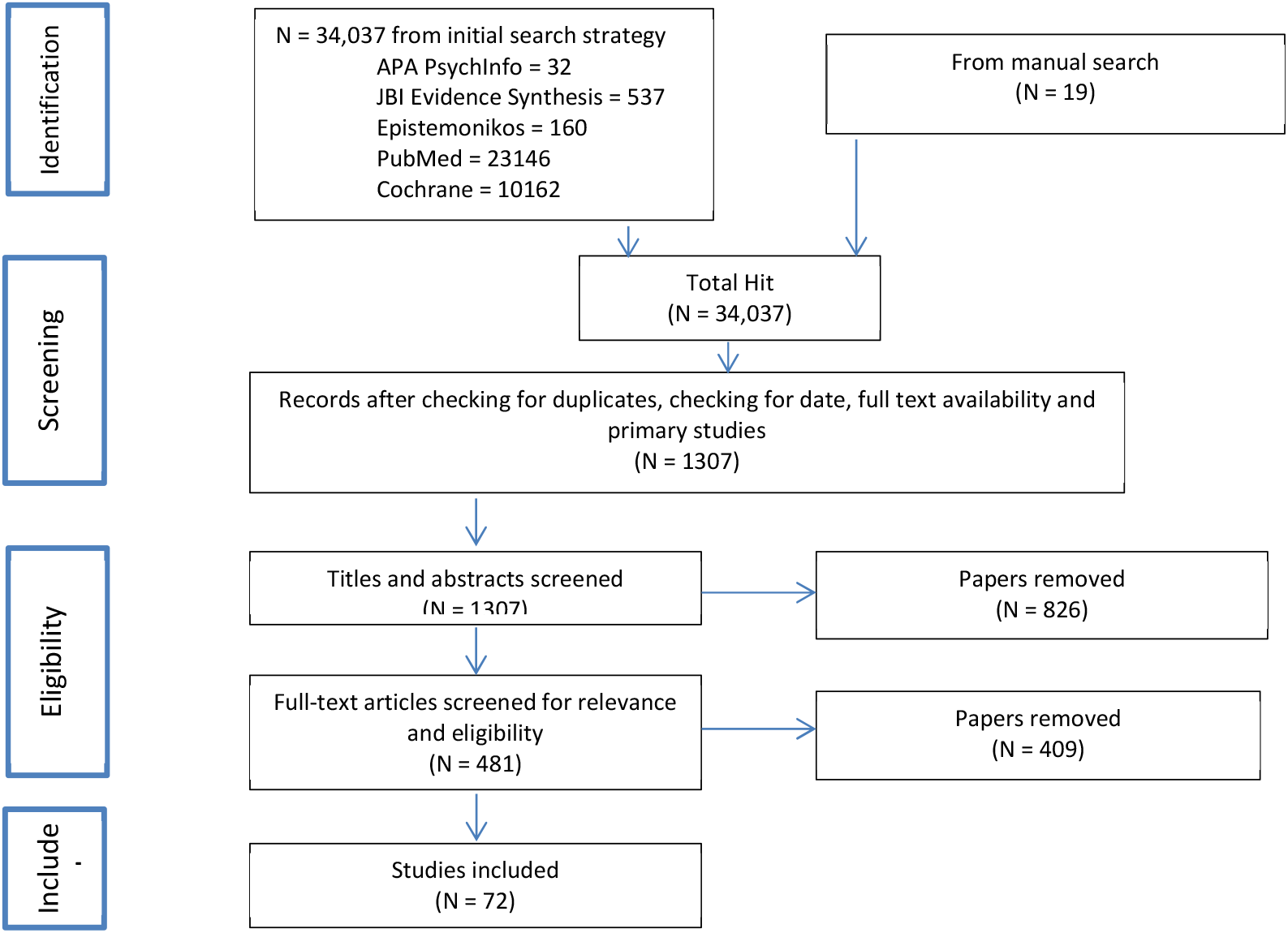
Flow diagram showing study selection results following PRISMA Recommendations

### Inclusion and exclusion criteria

The eligibility criteria for this review considered:

- peer-reviewed papers that focused on the mental health impact of the COVID-19 pandemic.
- primary research studies whose full texts were publicly and freely accessible.
- papers published after March 11, 2020 - when the WHO declared COVID-19 a pandemic.
- papers that were published in the English language.

### Data extraction, analysis, synthesis, and reporting

After a thorough full-text assessment and collation of 72 selected relevant articles, data extraction and setting up of the selected bibliography and abstracts was done using Mendeley Cite^®^ software. The abstracts were then examined for key findings which were then charted into a summary table (Table 2). Each study was then critically read to capture more information from the full texts. The findings were collated in Microsoft word documents and then key data was carefully transferred into Excel spreadsheets for further descriptive analysis. Results were then presented in tables and graphs. The information captured author names, country of study, study setting and population, study design, study aims and objectives, mental health assessment tools used, type, risk factors, and prevalence of mental health conditions studied. Where quantitative measurements were used, the numeric data were collated, grouped, and compared according to selected populations and geographical metrics such as gender, age, continents, and countries, and then presented in tables and chats. The risks identified were further summarised in a table (Table 5) and discussed as they relate to the research objectives. An arithmetic mean value was calculated for the prevalence of mental health conditions by taking the arithmetic mean of all the prevalence reported to provide an estimate of the trends of prevalence over time. Various gaps identified were further discussed with necessary recommendations.

**Table 2:** Data Chart Showing Study Characteristics.

| Author year | Study Design | Study Aim | Country, Sample size & characteristics, | Mental Health (MH) Condition Studied & Assessment tool used | Summary of Findings (Prevalence<sup>#</sup>) | Associated Risk factors |
| --- | --- | --- | --- | --- | --- | --- |
| He et al. (22) | CS | To assess the differences in mental health conditions among the general population, quarantined population, and healthcare workers during the COVID-19 outbreak in China | China; N = 2689 (374 general population, 403 healthcare workers, 1912 quarantined population, Adults ≥18). | PHQ-9 (depressive symptoms)<br>GAD-7 (anxiety) | Quarantined: Dep: 58.6%, anxiety: 41.2<br>Gen. Pop.: dep: 25.1%, anxiety: 18.5<br>HC workers: dep: 48.6%, anxiety: 35.7 | LRA:<br>All: Perceived effect on daily life, concerns posted on social media led to more depression and anxiety |
| Gramaglia et al. (23) | CS | To assess burnout, depression, anxiety, and PTSD symptoms in the ICU staff during the pandemic period | Italy; N = 95 (HCW of ICU in an Italian hospital, online survey participants, 67% female). | MBI (occupational burnout),<br>GHQ-12 (MH issues),<br>IES (PTSD),<br>BAI (anxiety),<br>BDI-II (depression) | Dep: 20%<br>Anxiety: 12%<br>PTS symptoms: 50% | Pearson's chi-square test, Fisher's, and Wilcoxon-type tests:<br>Increased mental health conditions were associated with gender (female), negative test results, and those who changed family habits because of the pandemic.<br>Influenced gen. MH problems.<br>Single or divorced or not cohabiting with partner influenced dep. symptoms,<br>Lack of previous ICU experience led to more anxiety |
| Gloster et al. (24) | CS | To determine mental health outcomes during pandemic-induced lockdowns and examine known predictors of mental health outcomes | International; N = 9565 (≥18 years, 78 countries, online survey, Latvia & Italy = 2242). | PSS (stress),<br>MSBS (depression),<br>PANAS (affect),<br>MHC-SF (wellbeing) | Low mental health: 10%<br>Moderate mental health: 50% | ANOVA, Univariable Linear regression, Linear mixed-effect model:<br>Increased risk of mental health conditions associated with lower social support level, lower education level, low psychological flexibility, lower financial status, less access to basic supplies |
| Gao et al. (25) | CS | To evaluate the mental health of nursing students during the COVID-19 pandemic | Finland; N = 1532 (nursing students in vocational college, average age: 19.95, 88% females) | IES-R (PTSD),<br>DASS-21 (depression, anxiety, stress),<br>PSQI (insomnia) | PTSD: 44.5%, Depression: 2.9%,<br>Anxiety: 2.9%, stress = 1.1%<br>Psychological interventions should be implemented for nursing students | Mann-Whitney U-test:<br>Higher risks of PTSD, depression, and anxiety among senior nursing students, gender (male), and urban students |
| Priyantini et al. (26) | CS | To analyze the factors that influence the MH crisis of COVID-19 infection on the island of Java | Indonesia; N = 1218 (69% women, 44.7% between 17-25 years | Brief-COPE (coping ability)<br>ADQ (MH crisis emergencies questionnaire, depression, anxiety, stress) | Heavy stress levels: 11.7%<br>Ver high level of anxiety: 23.6%<br>23.4% experiencing a Mental Health crisis | Chi-square test & Logistic Regression:<br>Mental health conditions associated with marital status (single), location (West Java), religious affiliation (Confucian), low-income range, lower coping mechanisms, severe stress levels |
| Davis et al. (27) | CS | To assess the perceived ability to comply with national COVID-19 mitigation strategies and their impact on household finances, food security, and mental health of medical and pharmacy students in Liberia | Liberia; N = 113 ($\geq 18$, medical and pharmacy students, 61.9% male, 75.2% single). | PHQ-8 (depression) | Positive depression screen (PHQ-9 $\geq 10$): 19.4% | Chi-square test, Mann-Whitney-Wilcoxon tests: Increased risk of depression associated with gender (male), living with a partner, concern about finances, concern about household's health |
| Chi et al. (28) | CS | To investigate the prevalence and risk factors for poor MH of Chinese university students during the COVID-19 pandemic | China; N = 2038 (students from 180 Chinese universities, avg. age: 20.6, 63% female). | PHQ-9 (depression), Z-SAS (anxiety), CD-RISC (resilience) | PTSD: 30.8%, anxiety: 15.5%, depressive symptoms: 23.3% | RIDGE regression: PTSD, depression, and anxiety associated with older age, knowing people who have been isolated, having more adverse childhood experiences, lower resilience |
| Chen and Li (29) | CS | To investigate the prevalence and influencing factors of anxiety, depression, perceived stress, and acute stress disorder among dental medical staff in emergencies during COVID-19 | China; N = 808 ($\geq 18$ years, avg. age: 36.2, emergency dental staff). | GAD-7 (anxiety), PHQ-9 (depression), PSS-10 (perceived stress), ASDS (acute stress) | Depression: 46.4%, anxiety: 36.3%, perceived stress: 65.2%, ASD: 1.1% | Chi-square test, non-parametric Mann-Whitney U-test, Kruskal-Wallis test: More anxiety is linked to Lower education level, Location (Wuhan) More perceived stress: previous medical history, Location (Wuhan) |
| Cai et al. (30) | CS | To investigate the psychological abnormality in HC workers battling the COVID-19 epidemic and to explore the associations among social support, resilience, and MH | China; N = 1521 (HC workers, 147 experienced staff, 1374 fresh staff) | SCL-90 (depression, anxiety), CD-RISC (resilience) | Psychological abnormality: 14.1% | Chi-square, t-test, multiple regression analyses: Increased risk of psychological abnormality associated with: Lower PH emergency experience, fresh staff |
| Buonsenso et al. (31) | CS | To ascertain the clinical characteristics, activity levels, and MH problems in children with long COVID | UK and USA; N = 510 (children with long COVID, avg. age: 10.3, 56.3% female). | LCKRS-2 (MH status) | Lack of concentration: 60.6%, Memory problems: 45.9%, Difficulty processing info.: 32.7%, Short-term memory issues: 32.7% Had 3 or more MH issues: 54.7% | ANOVA: Mental health conditions influencing factors: lockdown, school closure |
| Bettinsoli et al. (32) | CS | To investigate the psychological health of Italian HC professionals during the COVID-19 outbreak | Italy; N = 580 (HC workers in Italy, 59% male). | ADQ (psychological states), GHQ-12 (mental health status), CSES-brief (coping ability), BRCS (resilience) | 33.5% at risk for psychiatric morbidity | Chi-square, t-tests, linear regression: Increased risk of mental health conditions: occupation type (nurses), Gender (female) |
| Bella Nichole and Jonathan (33) | CS | To determine the presence of, and magnitude of associations between baseline associations and anxiety and | United States; N = 1005 (avg. age: 45, 48.8% male). | GAD-7 (anxiety), PHQ-9 (depression), ULS-8 (loneliness) | Anxiety disorder: 26.8% (cut-off of 10), clinical anxiety: 41.4% Depression: 21.6% | Logistic regression analyses: Increased risk of anxiety linked to gender (female), living in a smaller home, rural location, loneliness, history of hospitalization |
|  |  | depression in the US general population |  |  |  | Increased depression risks associated with gender (female), less time outdoors, smaller home, losing a job, loneliness, history of hospitalization |
| Angelina et al. (34) | CS | To investigate the magnitude of COVID-19-related MH problems in adolescents and the associated factors | Indonesia; N = 2018 (adolescents, 91% male, avg. age: 19). | K-10 (psychological distress, i.e., depression, nervousness, etc.) | Psychological distress: 54.1% | Mann-Whitney U test, Kruskal-Wallis test, Logistic Regression:<br>Increased risk of psychological distress associated with bad dietary patterns, low sleep quality, lack of confidence, frequent arguments with parents, worry, chronic diseases, mental illnesses |
| Alshumrani et al. (35) | CS | To evaluate sleep quality and assess the psychological burden of the pandemic on COVID-19 patients and the general population | Saudi Arabia; N = 1091 (58.9% +ve for COVID, 61.1% male). | PSQI (Sleep quality), PHQ-9 (depression), AIS (insomnia) | Poor sleep quality: 66.1% (COVID patients), 72.8% (gen. pop.)<br>Insomnia: 50.5% (COVID patients), 58.5% (gen. pop.)<br>Depression: 39.5% (COVID patients), 70.1% (gen. pop.) | Chi-square test, t-tests:<br>Increased MH issues: increased age |
| AlAteeq et al. (36) | CS | To explore depression and anxiety levels among HC providers during the COVID-19 outbreak in Saudi Arabia | Saudi Arabia; N = 502 (HC providers, 68.1% males). | PHQ-9 (depression), GAD-7 (anxiety) | Depressive disorder: 55.2% (mild to severe),<br>Generalized anxiety disorder: 51.4% | Multivariate analysis:<br>Increased anxiety associated with gender (female), nurses<br>Increased depression and anxiety: age range (30 – 39) |
| Murphy et al. (10) | CS | To explore the impact of COVID-19 and restrictions to daily living because of social distancing within veterans with pre-existing MH difficulties | United Kingdom N = 275 (treatment-seeking military veterans, avg. age: 48.7, 94.9% male). | GHQ-12 (CMD, e.g., depression, anxiety, etc.), PCL-5 (PTSD), DAR-5 (anger), AUDIT (alcohol misuse) | Common MH issues 69.3% (GHQ-12 ≥ 4),<br>PTSD: 65% (PCL-5 = 34),<br>anger issues: 52.7% (DAR-5 ≥ 12),<br>alcohol misuse: 30% (AUDIT ≥ 8) | Regression analyses:<br>Increasing severity of CMD & PTSD: lack of social support, increasing number of stressors |
| Steward et al. (37) | CS | Impact of COVID-19 on the MH and physical activity of pharmacy students at the University of Zambia | Zambia; N = 273 (undergraduate pharmacy students, University of Zambia, 51.6% female). | GAD-7 (anxiety) | Mild, moderate, and severe anxiety: 34.4%, 24.9%, and 16.9% respectively (GAD-7 = 5-9, 10-14, 15-21 respectively) | Partial proportion odds regression model:<br>Increasing MH issues linked to reduced family care, lack of attention to MH, year of study (final year), reduced resting time, feeling helpless |
| Morniroli et al. (38) | CS | To determine the prevalence of anxiety symptoms in new mothers throughout their hospital stay during the COVID-19 pandemic | Italy; N = 109 (negative COVID-19 newly delivered mothers in Italian hospital). | STAI-Y (anxiety) | Anxiety because of the pandemic: 42% (STATE-A ≥ 40) | Binary logistic regression models:<br>Increased anxiety linked to their father's absence during the hospital stay |
| Lu et al. (39) | CS | To investigate the mental and psychological health of adolescents in China and its possible related factors | China; N = 795 (adolescents, 73.2% male, avg. age: 17) March 19 – 29, 2020 | SDS (depression)<br>SAS (anxiety) | Depression: 76.48% (SDS ≥ 50)<br>Anxiety: 33.08% (SAS ≥ 50) | Multiple regression analyses: |
|  |  |  |  |  |  | Increased depression linked to gender (female), anxiety, history of mental disorders, family and social contradictions, COVID-19 knowledge reserve<br>Increased anxiety linked to gender (female), depression, MH knowledge reserves, family, and social contradictions |
| Jiang et al. (40) | CS | To investigate the sleep quality and MH status of HC professionals during the outbreak of COVID-19 | China; N = 569 (HC professionals, avg. age: 34, 59.9% females). | PSQI (sleep disorders), GAD-7 (anxiety), PHQ-9 (depression) | Sleep disorders: 28.8% (PSQI > 7)<br>Anxiety: 33.2% (GAD-7 > 4)<br>Depression: 39.4% (PHQ-9 >4) | Chi-square, t-test, Pearson correlation:<br>Sleep disorders linked to the occupation (frontline workers), underlying disease<br>Increased anxiety and depression linked to the occupation (frontline workers), underlying diseases, sleep quality |
| Jakhar et al. (41) | Coh. | To investigate how the psychological health of HC professionals on COVID duty was different from those who were not directly in contact | India; N = 450 (doctors and nurses, avg. age:31.6, 52% females). | DASS-21 (depression, anxiety, stress) | Stress: 33.8%<br>Anxiety: 38.9%<br>Depression: 43.6% (non-exposed), | Chi-square, Pearson correlation, logistic regression:<br>Increased stress, depression, and anxiety: COVID-19-exposed individuals (double that of non-exposed individuals), |
| Maehl et al. (41) | CS | To investigate whether the pandemic affected primary care and health outcome of a disease management program for coronary artery disease (DMP-CAD) patients | Germany; N = 750 (DMP-CAD patients, avg. age: 73.3, 67.2% male). | PHQ-9 (depressive symptoms)<br>GAD-7 (anxiety) | Total PHQ-9 mean score: 4.2 ± 4.2<br>Total GAD-7 mean score: 3.9 ± 4.1 | Univariate ANOVA:<br>Increased risk of mental health conditions linked to gender (female), reduced family visits, less frequency of leaving homes |
| Ma et al. (42) | CS | To assess the MH problems associated factors among a large sample of college students during the COVID-19 outbreak in China | China; N = 746217 (students from 108 colleges, 55.6% female). | PHQ-9 (depressive symptoms)<br>GAD-7 (anxiety)<br>IES-6 (acute stress) | Acute stress: 34.9%<br>Depressive symptoms: 21.1%<br>Anxiety: 11% | Univariate and hierarchical logistic regression analyses:<br>Increased risk of MH problems linked to having infected friends/relatives,<br>2.13x more risk of acute stress: exposure to media<br>Increased anxiety and depressive symptoms: low perceived social support (5x more), year of study (senior year), prior MH problems |
| Lugito et al. (43) | CS | To investigate the impact of social media exposure during the COVID-19 pandemic on depression, anxiety, and stress | Indonesia; N = 220 (participants, 18 years and above, avg. age: 24, 55% female, 98.2% had no family member with COVID-19). | DASS-21 (depression, anxiety, stress) | Mild-moderate depression: 29.5%<br>Severe-extremely severe depression: 46.4%<br>Mild-moderate anxiety: 19.6%<br>Severe-extremely severe anxiety: 60.9%<br>Mild-moderate stress: 16.3%<br>Severe-extremely severe stress: 38.2% | Pearson chi-square and multinomial logistic regression:<br>Severe-extremely severe anxiety: gender (female)<br>Mild-moderate depression: gender (female)<br>Reduced likelihood to suffer severe-extremely severe depression, mild-moderate, severe-extremely severe anxiety, and mild-moderate stress linked to increased social media exposure |
| S. Liu et al. (44) | CS | To investigate the prevalence and gender differences of various MH | China; N = 1563 (HC workers, 82.7% females). | PHQ-9 (depressive symptoms) | Depressive symptoms: 50.7% (PHQ-9 ≥ 5), | Mann-Whitney U-test, Pearson's chi-square test: |
| | | problems among HC workers in China during the COVID-19 outbreak | | GAD-7 (anxiety)<br>IES-R (stress)<br>ISI (insomnia) | Anxiety: 44.7% (GAD-7 $\geq 5$),<br>Stress: 52.5% (IES-R $\geq 20$),<br>Insomnia symptoms: 36.1% (ISI $\geq 8$) | Increased prevalence of all MH problems: gender (female), low living situations in the past week, location (Hubei province) |
| Liang et al. (45) | CS | To evaluate psychological symptoms in frontline medical workers during the COVID-19 epidemic in China in comparison with the general population | China; N = 1913 (899 frontline medical workers, 1104 general population respondents). | PHQ-9 (depression)<br>GAD-7 (anxiety)<br>ISI (insomnia)<br>CD-RISC-10 (resilience) | Depression in frontline workers: 30.43% (Hubei), 23.13% (others)<br>Depression in gen. pop.: 23.33% (Hubei), 18.25% (others)<br>Anxiety in frontline workers: 20.29% (Hubei), 13.14% (others)<br>Anxiety in gen. pop.: 16.67% (Hubei), 9.22% (others)<br>Insomnia in frontline workers: 14.9% (Hubei), 10.64% (others)<br>Insomnia in gen. pop.: 6.67% (Hubei), 7.17% (others) | Bonferroni's Post Hoc Multiple Comparison Test:<br>Increased resilience: Frontline staff in Hubei<br>Order of increasing MH issues: gen. population in other regions < gen. pop. in Hubei < medical workers in other regions < medical workers in Hubei province |
| Le et al. (46) | CS | To examine multiple types of COVID-related discrimination and their associations with MH outcomes among racial/ethnic groups in the US | United States; N = 1688 (adults, 44.3% non-Hispanic White, 19.9% prior mental health diagnosis). | PHQ-4 (psychological distress) | Psychological distress: 20.8% (PHQ-4 $\geq 6$) | Bivariable comparisons and adjusted logistic regression model:<br>Increased MH issues: perceived discrimination |
| Lawson et al. (47) | CS | To evaluate perceptions of delayed fertility care secondary to the COVID-19 pandemic | United States: N = 787 (patients from a single academic fertility center, participants were randomized 1:1 to receive education regarding fertility treatment, 82% female). | PHQ-8 (depression)<br>GAD-7 (anxiety) | Moderate to severe anxiety: 69.1% (education group), 73.7% (no education group)<br>Depression: 77.5% (education group), 77.9% (no education group) | Ordinal logistic regression:<br>Distress: related to age, duration of infertility, social support availability |
| Lai et al. (48) | CS | To investigate factors associated with MH outcomes among HC workers in China | China; N = 1257 (HC workers treating patients with COVID-19 in 34 hospitals, 64.7% aged 26 to 40, 76.7% females). | PHQ-9 (depression)<br>GAD-7 (anxiety)<br>ISI (insomnia)<br>IES-R (distress) | Depression: 50.4%<br>Anxiety: 44.6%<br>Insomnia: 34%<br>Distress: 71.5% | Multivariable logistic regression analysis:<br>Increased risk of distress: location (Wuhan)<br>Increased risk of depression, anxiety, insomnia, distress: frontline HC workers<br>Risk of severe symptoms of depression, anxiety, distress: gender (female) |
| Ko et al. (49) | CS | To evaluate traumatic stress and MH problems associated with the prolonged COVID-19 pandemic and to determine differences across age groups | Korea; N = 1151 (children/adolescents (19.9%), adults (56.6%), elderly (23.5%), avg. age: 41.03, 67% female). | PHQ-9 (depression)<br>CDI (depression in children)<br>GAD-7 (anxiety)<br>PSWQ-C (anxiety in children) | PTSD symptoms: 24.1% (gen.), 14.4% (children/adolescents), 30.8% (adults), 15.9% (elderly) (PC-PTSD-5 $\geq 3$)<br>Depression: 20.9% (gen.)<br>4.4% (children/adolescents), | Chi-square analysis, Pearson's correlation analyses:<br>Increased risk of all MH issues: highest in the adult group<br>In children and adolescents, increased depression led to increased PTSD symptoms |
| | | | | P4 screener (suicide risk)<br>PC-PTSD-5 (PTSD symptoms) | 28.8% (adults), 15.9% (elderly) (PHQ-9 ≥ 10, CDI ≥ 26)<br>Anxiety: 16.8% (total)<br>7.4% (children/adolescent), 22.5% (adult), 10.7% (elderly) (GAD-7 ≥ 10, PSWQ-C ≥ 21)<br>Suicide risk: 20.5% (total)<br>20.9% (adolescent group), 25% (adult group), 9.3% (elderly group) | In both the adult and elderly groups, depression is positively related to an increased risk of PTSD symptoms |
| Katz et al. (50) | CS | To examine changes in symptoms of anxiety and depression from pre-pandemic to during the pandemic | United States; N = 1504 (avg. age: 66.1, 85.7% female). | PHQ-4 (anxiety and depression) | Increase in anxiety: 10.6%<br>Increase in depression: 7.9% | Multiple logistic regression:<br>COVID-specific stress ratings = 2x gen. stress ratings<br>Increased depression and anxiety: general and COVID-19-specific stress |
| Kalita et al. (51) | Coh | To evaluate the impact of COVID-19 and lockdown on the physical and MH, and quality of sleep in a cohort of myasthenia gravis patients | India; N = 38 (median age: 45, 57.9% female). | HADS (anxiety and depression)<br>PSQI (sleep quality) | Depression: 5% (HADS-D > 11)<br>The median quality of sleep reduced from 4 to 3<br>The Median HADS score increased from 2 to 3 | Mann Whitney U test, chi-square test:<br>Depression, anxiety, and quality of sleep were worsened for participants as a result of the pandemic |
| Kang et al. (48) | CS | To explore the MH status of medical and nursing staff and the efficacy of critically connecting psychological needs to receiving psychological care | China: N = 994 (medical and nursing staff in Wuhan, 85.5% female, 63.4% aged 25 – 40, participants were grouped into 4 groups of “disturbances”). | PHQ-9 (depression)<br>GAD-7 (anxiety)<br>ISI (insomnia)<br>IES-R (distress) | MH disturbances: 36.9%<br>Mild disturbances: 34.4%<br>Moderate disturbances: 22.4%<br>Severe disturbances: 6.2% | Chi-square test, structural equation model:<br>Increased disturbances: gender (female)<br>Groups with higher distress had a more extensive scope of exposure |
| Hu et al. (52) | CS | To assess the prevalence of both depression and anxiety symptoms among civil servants in Jiangsu and surrounding provinces | China: N = 867 (Chinese civil servants aiding in COVID-19 control, 18 years and above). | PHQ-9 (depression)<br>GAD-7 (anxiety) | Depression: 37.25%<br>Anxiety: 38.06% | Multivariable logistic regression analysis:<br>Increased depression and anxiety: younger age, more education, less work experience, gender (female) |
| Islam et al. (21) | CS | To assess the prevalence of depression, anxiety and stress, and associated factors among university students in Bangladesh | Bangladesh; N = 3122 (Bangladeshi university students, 59.5% males, avg. age: 21.4). | DASS-21 (depression, anxiety, stress) | Depression: 76.1% (mild to severe)<br>Anxiety: 71.5% (mild to severe)<br>Stress: 70.1% (mild to severe) | Regression analyses:<br>Increased DAS: lower students’ satisfaction with studies during the pandemic |
| Htun et al. (53) | CS | To assess the prevalence and associated factors of depressive symptoms in patients with COVID-19 at the Hmawbi treatment center | India; N = 142 (patients with COVID-19 at Hmawbi Treatment Centre). | CES-D (depressive symptoms) | Depressive symptoms: 38.7% | Bivariable logistic regression analysis:<br>Increased depressive symptoms: 40 years and above, household size less than 4, lower monthly family income, infection to family members |
| Hou et al. (54) | CS | To examine the effect of social support on MH of HC workers and its underlying mechanisms | China; N = 1472 (HC workers, 76.5% female). | CD-RISC (resilience)<br>SCL-90 (mental health evaluation) | Psychological abnormality: 7% (SCL-90 ≥ 160) | Structural equation modeling:<br>Resilience was positively linked to SCL-90 scores |
| Shen et al. (55) | CS | To explore the psychological status of medical staff in the epidemic period of COVID-19 and to analyze its influential factors | China; N = 373 (medical staff room 2 Chinese hospitals, 87% female, avg. age: 29.55). | SCL-90 (MH disorders) | Terror symptoms: 11.26%<br>Compulsive symptoms: 7.77%<br>Anxiety: 5.63% | Stepwise regression analysis:<br>The mental health of medical staff was influenced by the impact of COVID-19 information on their life, recent cough or fever, insomnia, fear of infection |
| Shah et al. (5) | CS | To assess the impact of the COVID-19 pandemic and lockdown on MH symptoms in children | India; N = 423 (children aged 11-15, 54.4% male). | PSC (MH disorders) | Psychosocial problems: 30.7%<br>Anxiety or depressive symptoms: 25.2% | Binary regression analysis:<br>Increased psychosocial problems: fear of COVID-19 infection, inability to attend school, distance from friends<br>Higher risk of anxiety or depressive symptoms: increased social media use |
| Severinsen et al. (56) | Coh | To compare the MH response among pregnant women with that in similarly aged women from the general population during the first wave of the COVID-19 pandemic | Denmark; N = 1505 (647 pregnant women in their 2<sup>nd</sup> trimester, 858 women from the general population, the median age for pregnancy Women: 31.4, the median age for gen. pop.: 35). | CMHDQA-4 (anxiety)<br>ULS-3 (loneliness)<br>CCMH (MH disorders) | 70.3% negative feelings (pregnant women) | Non-parametric Wilcoxon rank sum tests, chi-square tests:<br>Pregnant women were more concerned about becoming sick but less lonely<br>Pregnant women reported more negative feelings |
| Ruengorn et al. (57) | CS | To evaluate the association of economic burden during the first phase of the pandemic and the risk of adverse MH outcomes in the Thai population | Thailand; N = 2303 (18 years and above, 60% females, avg. age: 34.5) April 21 – May 4, 2020 | PHQ-9 (depression)<br>GAD-7 (anxiety)<br>PSS-10 (perceived stress) | Job loss led to: 51.9% depressive symptoms (PHQ-9 ≥ 9), 56.9% anxiety (GAD-7 ≥ 5), 84.7% perceived stress (PSS ≥ 14)<br>≥ 50% income loss led to: 43.2% depressive symptoms, 49.4% anxiety, 78.4% perceived stress<br>Financial problems led to 43.5%, 50.4%, and 80.2% depressive, anxiety, and perceived stress symptoms respectively. | Multivariable logistic regression models<br>Increased risk of perceived stress: loss of job because of COVID-19<br>Increased risk of anxiety: loss of 50% of monthly income or more<br>MH issues: self-reported financial problems |
| Pieh, Budimir and Probst (58) | CS | To evaluate MH during the COVID-19 lockdown in Austria | Austria; N = 1005 (53% female). | PHQ-9 (depressive symptoms)<br>GAD-7 (anxiety)<br>ISI (insomnia)<br>PSS-10 (Perceived stress) | Depressive symptoms: 21% (PHQ-9 ≥ 10)<br>Moderate anxiety symptoms: 19% (GAD-7 ≥ 10)<br>Clinical insomnia: 16% (ISI ≥ 15) | Bonferroni-corrected posthoc tests, t-tests:<br>Highest MH problems: adults under 35, gender (female), people with no work, low income |
| Qin et al. (59) | CS | To investigate the MH of patients with COVID-19 in Changsha, China | China: N = 112 (patients with COVID-19, $\geq 18$ years, 52.7% male, median age: 40). | SCL-90 (MH issues) | MH issues 21.4% (SCL-90 $\geq 160$) | Mann-Whitney U test, chi-square test, t-tests:<br>Increased MH symptoms: less sleep time, gender (female) |
| Rossi et al. (60) | CS | To investigate MH outcomes among frontline and second-line HC workers during the COVID-19 pandemic in Italy | Italy: N = 1379 (HC workers, 77.2% female, avg. age: 39). | GPS (posttraumatic stress symptoms)<br>PHQ-9 (depression)<br>GAD-7 (anxiety)<br>ISI (insomnia)<br>PSS (perceived stress) | PTSD: 49.38% (GPS $\geq 3$)<br>Depressive symptoms: 24.73% (PHQ-9 $\geq 15$)<br>Anxiety: 19.8% (GAD-7 $\geq 15$)<br>Insomnia: 8.27% (ISI $\geq 22$)<br>High perceived stress: 21.9% | Multivariable logistic regression model:<br>Increased PTSS, depression, anxiety, perceived stress: linked to gender (female), younger age, having a deceased colleague or colleague in quarantine<br>Increased PTSS, depression, insomnia:<br>Increased insomnia: occupation (nurses, HC assistants) |
| Prati (61) | CS | To investigate the psychological impact of national quarantine in Italy | Italy: N = 1569 (81.5% female, avg. age: 31.3). | GHQ-12 (MH symptoms) | Common MH disorders: 31.7% (men), 52.3% (women) | ANCOVA:<br>Increased MH issues: gender (female), lower age, worry, lower media, employed, lower coping efficacy, lower trust in institutions |
| Penteado et al., (62) | CS | To examine the magnitude of psychiatric symptoms and to determine their association with caregiver distress | Brazil: N = 100 (71 older adults with psychogeriatric/neurocognitive disorders, 29 aging adults with down syndrome). | HADS (anxiety, depression)<br>NPI-Q (psychological and behavioral symptoms) | Depression: 50.7% (elderly), 3.4% (others)<br>Neurocognitive disorder: 90.1% (elderly), 34.4% (others)<br>Anxiety: 65% (elderly) | ANOVA, linear regressions:<br>Higher HADS and NPI-Q scores were linked to caregiver distress in all groups<br>Higher HADS scores: patients with dementia |
| Norhayati, Che Yusof and Azman (63) | CS | To compare the levels of vicarious traumatization between frontline and non-frontline HC providers in response to the COVID-19 pandemic | Malaysia: N = 306 (160 frontline, 146 non-frontline HC providers). | VTQ (vicarious traumatization) | Mean Vicarious traumatization: 79.7 (non-frontline)<br>Mean vicarious traumatization: 74.3 (frontline) | ANCOVA:<br>Higher vicarious traumatization score: gender (female), lower social support |
| Nguyen et al. (64) | CS | To explore COVID-19-related, psychological stress risk factors among HC workers, their concerns, and demands for MH support during the pandemic | Vietnam: N = 761 (HC workers, 58.2%) | IES-R (psychological response to trauma) | Stress: 34.3% | Chi-square test, Fisher's exact test:<br>Increased risk of stress linked to being a frontline HC worker, perceived worse well-being compared to before the pandemic, suffering chronic diseases |
| Naser et al. (65) | CS | To explore the prevalence of depression and anxiety among the general population, HC practitioners, and university students during the COVID-19 outbreak | Jordan: N = 4126 (1798 gen. pop., 1163 HC practitioners, 1165 students; 59% female, 53.9% single). | PHQ-9 (depression)<br>GAD-7 (anxiety) | Depression: 23.8% (PHQ-9 $\geq 15$)<br>Anxiety: 13.1% (GAD-7 $\geq 15$) | Logistic regression analysis:<br>Increased depression linked to divorced, pulmonologists, students with chronic diseases, female HC professionals, and students<br>Increased anxiety risk linked to gender (female), divorced (gen. pop.), university students with chronic diseases, high income |
| Shi et al. (66) | CS | To investigate the prevalence of and risk factors associated with MH symptoms in the gen. pop. in China during the COVID-19 pandemic | China: N = 56679 (avg. age: 35.97, 52.1% female). | PHQ-9 (depression)<br>GAD-7 (anxiety)<br>ISI (insomnia)<br>ASDS (stress) | Depression: 27.9%<br>Anxiety: 31.6%<br>Insomnia: 29.2%<br>Acute stress: 24.4% | Logistic regression analyses:<br>Increased risk of depressive symptoms, anxiety, acute stress, and insomnia linked to having family or friends with confirmed or suspected COVID-19; residence |
|  |  |  |  |  |  | (Hubei), having occupational exposure risks; being quarantined<br>Increased risk of depression, anxiety, and insomnia linked to not working |
| Simegn et al. (67) | CS | To assess depression, anxiety, and stress and identify associated factors among university students in Ethiopia during the early stage of the COVID-19 pandemic | Ethiopia: N = 423 (university students, 64.3% male, avg. age: 22.96). | DASS-21 (depression, anxiety, stress) | Depression: 46.3%<br>Anxiety: 52%<br>Stress: 28.6% | Binary logistic regression:<br>Increased risk of depression: lack of knowledge, study materials, and access to internet resources about COVID-19 prevention.<br>Increased risk of anxiety is linked to gender (female), lower ages, non-health-related students, thinking COVID-19 is unpreventable, lack of information about COVID-19 prevention<br>Increased risk of stress: linked to gender (female), students in 1<sup>st</sup> and 2<sup>nd</sup> years, thinking COVID-19 is unpreventable, COVID-19 patients in their town of residence, lack of study materials |
| Xingyue et al. (68) | CS | To assess the MH of emergency department medical staff during the epidemic in China | China: N = 14825 (doctors and nurses). | CES-D (depressive symptoms)<br>PCL-5 (PTSD) | Depressive symptoms: 25.2% (CES-D ≥ 16)<br>PTSD: 9.1% | Logistic regression models:<br>Increased risk of depressive symptoms and PTSD: gender (males), age (middle-aged), worked fewer years, worked for long hours daily, lower social support<br>Increased risk of depressive symptoms: work location (Hubei)<br>Increased risk of PTSD: occupation (nurse) |
| Teixeira et al. (69) | CS | To verify the prevalence of symptoms of psychological distress in medical students during the COVID-19 pandemic | Brazil: N = 656 (medical students). | SRQ-20 (psychological distress) | Psychological distress: 62.8% (SRQ-20 ≥ 7) | Pearson's chi-square test:<br>Increased risk linked to gender (female), being in 1<sup>st</sup> or 2<sup>nd</sup> year of study, struggling with adapting to distance learning, difficulty concentrating, worry about delay in graduation, previous mental disorder diagnosis, inability to maintain healthy habits, fear of infection |
| Zheng et al. (70) | CS | To assess the sleep quality, MH status, and associated factors among medical workers during the COVID-19 pandemic | China: N = 207 (medical workers, 84.54% female, 63.29% > 30 years). | PSQI (sleep quality)<br>SCL-90 (MH status) | Poor sleep quality: 34.3% (PSQI > 10)<br>MH symptoms: 27.05% (GSI > 1.50) | Logistic regression analysis:<br>Increased risk of poor sleep quality linked to gender (male), work experience > 15 years, profession (nurse), more night shifts, supporting Wuhan |
| Zhang et al. (71) | CS | To investigate and compare the prevalence and severity of MH symptoms between frontline medical staff and non-frontline medical staff during the COVID-19 outbreak | China: N = 524 (150 frontlines, 374 non-frontline medical staff). | ADQ (occupational stress)<br>GAD-7 (anxiety)<br>PHQ-9 (depression)<br>ISI (insomnia) | Depression: 31.3%<br>Anxiety: 41.2%<br>Insomnia: 39.3% | Hierarchical multiple regression models:<br>Increased levels of anxiety, depression, and insomnia: a medical (especially frontline) staff<br>Increased risk of MH issues: gender (male), marital status (married), poorer physical health, higher occupational stress (in all participants) |
| G. Y. Zhang et al. (72) | CS | To observe the impact of the COVID-19 pandemic on MH outcomes among patients with mild to moderate illness in Fangcan shelter hospitals | China: N = 129 (patients, avg. age: 43.8 years, 62% male). | SRSS (sleep condition)<br>PHQ-9 (depressive symptoms)<br>GAD-7 (anxiety symptoms) | Depressive symptoms: 49.5% (PHQ-9 ≥ 10)<br>Insomnia: 14.7% (SRSS ≥ 23)<br>Anxiety symptoms: 49.6% (GAD-7 ≥ 7) | Correlation analysis:<br>Increased depressive and anxiety symptoms linked to sleep disturbances, hypochondriasis<br>Depression, anxiety, distress: gender (female)<br>The COVID-19 pandemic was linked to depressive, and anxiety symptoms, sleep disturbances, and hypochondriasis; it also led to lower psychological resilience |
| Ying Zhang et al. (73) | CS | To investigate the incidence and risk factors of MH status among Infection Control Professionals (ICPs) in China during the COVID-19 outbreak | China: N = 9228 (ICPs from 3776 hospitals, 93.25% female, avg. age: 42.26, 80% nurses). | GHQ-12 (MH status)<br>PCQ (psychological capital) | MH problems: 60.77% (GHQ-12 ≥ 3)<br>Poor psychological capital: 39.2% | Chi-square test, backward stepwise regression:<br>Increased risk of MH problems linked to the place of work (public hospital), greater self-efficacy<br>Reduced risk of MH problems linked to marital status (unmarried), working years ≤ 1 year, fewer working hours per week, feeling hopeful/optimistic |
| Zhang et al. (74) | CS | To assess the psychological status of HC workers who were exposed to different risk levels in China and explore the factors that affected their MH | China: N = 810 (participants, 70.5% nurses, 81.7% female). | FSS (fatigue)<br>GAD-7 (anxiety)<br>ISI (insomnia) | Fatigue: 74.3% (FSS ≥ 36)<br>Anxiety: 73.7%<br>Insomnia: 61.7% | Binary logistic regression analysis:<br>Increased risk of MH problems: profession (nurses), gender (female), workers exposed to high-risk areas<br>Increased symptoms of fatigue, anxiety, and insomnia linked to exposure to high-risk areas |
| Zhan et al. (75) | CS | To understand medical students' MH during the COVID-19 pandemic | China: N = 266 (medical students from 4 schools). | DASS-21 (depression, anxiety, stress) | Depression: 20.3%<br>Anxiety: 17.7%<br>Stress: 6.4% | ANOVA, chi-square test:<br>Increased depression found among master's students than among college students, nursing majors |
| Ying et al. (76) | CS | To investigate the MH status and related factors in families of HC workers employed in designated hospitals in Ningbo, China | China: N = 845 (family members of HC workers, 52.66% male, median age: 37 years). | GAD-7 (anxiety)<br>PHQ-9 (Depression) | Anxiety: 33.73% (GAD-7 ≥ 5)<br>Depression: 29.35% (PGQ-9 ≥ 5) | Multivariable logistic regression analyses:<br>Increased risk of anxiety linked to thinking longer about COVID-19, a family member with direct contact with COVID-19 patients<br>Increased depression linked to thinking longer about COVID-19, longer work time per week for Healthcare family members, having a Healthcare worker as a parent or next of kin, private sector workers |
| Xiaoxv et al. (77) | CS | To quantitatively evaluate the influence of the COVID-19 outbreak on residents' MH | China: N = 8151 (residents, avg. age: 31.7). | PHQ-9 (depression)<br>GAD-7 (anxiety) | Depressive symptoms: 15.02%<br>Anxiety symptoms: 7.69%<br>Depressive and anxiety symptoms increased by 11% and 15% respectively between February 2019 and February 2020 | Multivariable logistic regression analysis:<br>Increased anxiety and depressive symptoms related to self-reported health status, exercise, social support, perceived death toll, treatment difficulty of COVID-19 |
| Yang et al. (78) | CS | To assess the impact of COVID-19 on the MH of university students in Sichuan, China | China: N = 521 (medical and non-medical students, avg. age: 22.02, 77.5% female). | SRQ-20 (distress)<br>SAS (anxiety) | Distress: 19% (SRQ-20 $>$ 7)<br>Mild, moderate, and severe anxiety: 31.5%, 8.1%, 5.8% | Logistic regression analysis:<br>Increased distress among respondents: ≤ 22 years |
|  |  |  |  |  |  | Increased risk of anxiety: respondents who paid attention to pandemic information, medical students |
| Xu et al. (79) | CS | To investigate the potential factors associated with MH outcomes among Chinese adults during the COVID-19 epidemic | China: N = 1456 (participants, avg. age: 33.8, 59.1% female). | ULS-3 (loneliness)<br>PHQ-2 (depression)<br>GAD-2 (anxiety) | Depressive symptoms: 11.3% (PHQ-2 $\geq 3$)<br>Anxiety symptoms: 7.6% (GAD-7 $\geq 3$)<br>Loneliness: 38.7% (ULS-3 $\geq 4$)<br>PTSD symptoms: 33.9% | Multivariable analysis:<br>Loneliness linked to marital status (single, separated/divorced/widowed, low education, other somatic symptoms, lower self-efficacy, going out frequently<br>Depression is associated with fear of infection, more somatic symptoms, lower self-efficacy, longer screen time, binge drinking<br>Anxiety is linked to more somatic symptoms, lower self-efficacy<br>PTSD symptoms are linked to more somatic symptoms, lower self-efficacy, increased perception of the risk of infection |
| Xie et al. (80) | CS | To investigate depressive and anxiety symptoms among students in Hubei province, China | China: N = 1784 (students, 845 from Wuhan, 1485 from Huangshi). | CDI-S (depressive symptoms)<br>SCARED (anxiety) | Depressive symptoms: 22.6%<br>Anxiety symptoms: 18.9% | Logistic regression analysis:<br>Increased risk of depressive symptoms, Higher CDI-S scores: students in Wuhan, were more worried about being affected by COVID-19, and students who were not optimistic about the epidemic |
| C. Wang et al. (81) | CS | To compare the levels of impact of COVID-19 on MH among people from Spain and China | China & Spain: N = 1528 (687 Spanish and 841 Chinese participants, avg age: 43.06 years (Spanish), 24.73 years (Chinese). | IES-R (MH impact)<br>DASS-21 (depression, anxiety, stress) | Spanish participants reported higher depression and stress scores<br>Chinese participants: higher IES-R scores | Linear regression, chi-square test:<br>Increased risk of adverse MH in Spain: more exposure to health information<br>Higher DASS-21 and IES-R scores were linked to headache, myalgia, and dizziness in both countries |
| Q. Wang et al. (82) | CS | To compare the MH and psychological responses in Wuhan and other areas of China during the COVID-19 epidemic | China: N = 4191 (1397 from Wuhan, 2794 from other areas). | PHQ-9 (depression)<br>ISI (insomnia) | MH issues: 46.6% (Wuhan), 32.2% (others)<br>Anxiety: 15.2% (Wuhan), 6.2% (others)<br>Depression: 18.3% (Wuhan), 9.7% (others)<br>Suicidal ideation: 10.5% (Wuhan), 7.1% (others)<br>Insomnia: 38.6% (Wuhan), 27.6% (others) | Regression analysis:<br>Living in Wuhan was linked to a higher rate of help-seeking behavior, feeling fearful, and increased risk of MH issues |
| Xia et al. (83) | CS | To investigate sleep quality and MH of patients with Parkinson's disease during the COVID-19 pandemic | China: N = 288 (119 PD patients, 169 healthy controls). | PSQI (sleep quality)<br>HADS (psychological distress) | Mean HADS score: 4.89% (PD patients), 3.82% (control)<br>Mean PSQI score: 8.13% (PD patients), 5.36% (control) | Logistic regression analysis:<br>Increased sleep disturbance in PD patients linked to exacerbation of PD symptoms, anxiety<br>Higher PSQI scores, anxiety, and depression in PD patients linked to gender (female) |
|  |  |  |  |  | Sleep disturbance: 68.9% (PD patients), 44.4% (others) |  |
| Wauters et al. (84) | CS | To investigate MH outcomes among parents of children with a chronic disease during the COVID-19 pandemic | Belgium: N = 1016 (599 parents of children with CD, 417 parents of healthy children). | PROMIS (anxiety, depression)<br>PSQI (sleep quality) | Mean Parental anxiety: 2.4 (clinical sample), 1.65 (control)<br>Mean parental depression: 1.7 (clinical sample), 1.59 (control) | Correlational and regression analyses:<br>Increased risk of parental anxiety: younger parents, mothers, parents of children with CD<br>Increased parental depression: younger parents, parents living alone, single parents |
| Wang et al. (85) | CS | To explore the psychological characteristics and influencing factors of college students after experiencing the impact of the epidemic and quarantine measures for epidemic prevention and control in the early stage of the outbreak of novel COVID-19 | China: N = 3641 (college students, 71.74% female, avg. age: 22.5 years). | SCSQ (coping ability)<br>DASS-21 (depression, anxiety, stress)<br>IES-6 (MH impact) | Negative emotions (depression, anxiety, stress, others): 14.69%<br>Depressive symptoms: 11.04%<br>Anxiety symptoms: 10.49%<br>Stress symptoms: 4.9%<br>PTSD: 34.19% | Logistic regression analysis:<br>Increased risk of depressive symptoms and PTSD: gender (male)<br>Decreased risk of depression, anxiety, stress, and PTSD: more family support |
| Teng, Wu and Xu (86) | CS | To investigate the psychological effects of operating a quarantined hotel on its employees | China: N = 170 (participants, 58.2% female, 53% aged 26 – 44 years). | DASS-21 (depression, stress, anxiety) | Depressive symptoms: 43.5%<br>Anxiety: 68.2%<br>Stress: 8.2% | ANOVA, regression analysis:<br>Increased anxiety: senior high school/vocational school employees (then in undergraduate and above), lower monthly incomes |
| Niekerk and van Gent (87) | CS | To determine the MH and well-being of staff members in an Eastern Cape university just after levels 4 and 5 lockdowns in South Africa | South Africa: N = 280 (staff members, avg. age: 48.84). | K-10 (psychological distress)<br>MHC-SF (psychological, emotional, and social well-being) | Psychological distress: 27.6%<br>Mental illness: 72.5% (K-10)<br>Moderate or low wellbeing: 39.3% | ANOVA:<br>Increased risk of psychological distress: younger age, gender (female), staff members with comorbidities, workers in administration and service sections<br>Higher mental well-being linked to increased age |
| Vujčić et al. (88) | CS | To determine the impact of the COVID-19 epidemic on MH of the general adult Serbian population and to identify associated factors during the state of emergency and lockdown | Serbia: N = 1057 (participants from 146 Serbian cities, avg. age: 36.06 years, 67.7% female). | DASS-21 (depression, anxiety, stress) | Severe depression: 28.9%<br>Anxiety: 36.9%<br>Stress: 38.1% | Multiple ordinal regression:<br>Increased risk of depression, anxiety, and stress linked to feeling of helplessness, perceived likeliness of impending death, experiencing COVID-19 symptoms<br>Increased risk of depression and stress linked to current smoking status, younger age, student population<br>Decreased risk of depression, anxiety, and stress linked to higher socioeconomic status |

CS = Cross-sectional Study, Coh = Cohort Study.
#Reports about the mental health impact of COVID-19 on the population under study.

### Quality appraisal

The Newcastle-Ottawa Scale (NOS) (18) was used to assess the methodological quality of primary studies in this review. For cross-sectional studies, we used a modified version of the NOS, as described by Modesti et al., (19). The modified NOS contains 3 major sections, with a total of 7 categories, which assess representativeness, sample size, non-respondents comparability, risk factor, confounding factors, assessment, and statistical issues. Quality assessment on cohort studies was carried out using a modified NOS for cohort studies (20). It has a total of 8 categories assessing representativeness, selection of non-exposed cohort, exposure, the outcome of interest, confounders control, outcome assessment, and follow-up duration and adequacy graded over 9 stars. The final quality scores for each study were assigned by modifying the scales used in previous studies (18–21): A score of 7 and above denoted a high-quality paper with low risk for bias, 4 – 6 were moderate quality papers with low risk for bias, and scores less than 4 were considered very low-quality papers with high risk for bias (18).

## Results

### Search results

The initial search of APA PsychInfo, JBI Evidence Synthesis, Epistemonikos, PubMed, and Cochrane databases produced 34,037 results (Table 1). Of these results, 2,016 were primary studies with the accessible full text, out of which 1,672 studies were published between March 2020 – July 2022. The references were initially imported into Mendeley reference management software, where 365 were identified as duplicates and removed, leaving a total of 1,307. The abstracts and titles were screened. A total of 826 papers did not meet the relevance and/or eligibility requirements for the review. The full text of the remaining 481 papers was assessed and 72 papers met the inclusion criteria. and their quality.

### Summary of studies

Most studies (95.8%) used a cross-sectional design methodology. A few studies (4.2%) were cohort studies. Three-quarters of the studies had more female participants than males. The total number of study participants was 914,078. Three papers studied multiple countries, but a greater number (32) of the studies were conducted in China (44%), 5 in Italy (7%), 4 each in the United States and India (6% each), 3 in Indonesia (4%), 3 each in Brazil and Saudi Arabia (3% each), and 1 from every other country (Table 3), making up a total of 24 countries spanning 5 continents; Africa, Asia, Europe, and North and South America. The study objectives varied slightly around the mental health of participants during the pandemic, a one-time comparison between different groups, or a time-lapse comparison of mental health statistics before and during the pandemic in a single group. A total of 27 papers were published in 2020, 34 in 2021, and 11 in 2022. Almost all the studies collected data directly in 2020 (68), and 2021 (4), but 5 studies did not report the exact time except for data submission/publication dates.

**Table 3:** Result of quality appraisal (cross-sectional studies: Max. Score = 10)

| Paper | Selection |  |  |  | Comparability | Outcome |  | Total score |
| --- | --- | --- | --- | --- | --- | --- | --- | --- |
|  | Representativeness | Sample size | Non-respondents | Ascertainment of exposure | Confounding factors | Outcome assessment | Statistical test |  |
| Vujčić et al., 2021 | * | * |  | ** | ** | * | * | 8 |
| Niekerk and van Gent, 2021 | * | * |  | ** | ** | * | * | 8 |
| Teng, Wu and Xu, 2021 |  | * |  | * | ** | * | * | 7 |
| C. Wang et al., 2021 | * | * |  | ** | ** | * | * | 8 |
| Wauters et al., 2022 |  | * |  | ** | ** | * | * | 7 |
| Xia et al., 2020 | * |  |  | ** | ** | * | * | 7 |
| Q. Wang et al., 2021 |  | * |  | ** | ** | * | * | 7 |
| Wang et al., 2022 |  | * | * | ** | ** | * | * | 8 |
| Xie et al., 2020 |  | * | * | ** | ** | * | * | 8 |
| Xu et al., 2021 |  | * |  | ** | ** | * | * | 8 |
| Yang et al., 2021 |  | * | * | ** | ** | * | * | 8 |
| Xiaoxv et al., 2020 | * | * |  | ** | ** | * | * | 8 |
| Ying et al., 2020 |  | * | * | ** | ** | * | * | 8 |
| Zhan et al., 2020 |  | * |  | ** | ** | * | * | 7 |
| Zhang et al., 2022 |  | * | * | ** | ** | * | * | 8 |
| Ying Zhang et al., 2021 | * | * |  | ** | ** | * | * | 8 |
| G. Y. Zhang et al., 2021 |  | * |  | ** | ** | * | * | 7 |
| Zhang et al., 2020 |  |  |  | ** | ** | * | * | 6 |
| Zheng et al., 2021 |  |  |  | ** | ** | * | * | 6 |
| Teixeira et al., 2021 |  |  |  | ** | ** | * | * | 6 |
| Xingyue et al., 2020 | * | * |  | ** | ** | * | * | 8 |
| Shi et al., 2020 |  | * | * | ** | ** | * | * | 8 |
| Simegn et al., 2021 | * | * | * | ** | ** | * | * | 9 |
| Naser et al., 2020 | * | * |  | ** | ** | * | * | 8 |
| Nguyen et al., 2021 | * |  |  | ** | ** | * | * | 7 |
| Norhayati, Che Yusof and Azman, 2021 |  | * | * | ** | ** | * | * | 8 |
| Penteado et al., 2020 | * | * |  | ** | ** | * | * | 8 |
| Prati, 2021a |  | * |  | ** | ** | * | * | 7 |
| Rossi et al., 2020 |  |  |  | ** | ** | * | * | 6 |
| Qin et al., 2020 |  |  |  | ** | * | * | * | 5 |
| Pieh, Budimir and Probst, 2020 | * | * |  | ** | ** | * | * | 8 |
| Ruengorn et al., 2021 |  | * |  | ** | ** | * | * | 7 |
| Shah et al., 2021 |  | * | * | ** | ** | * | * | 8 |
| Shen et al., 2020 |  | * | * | ** | ** | * | * | 8 |
| Hou et al., 2020 |  | * |  | ** | ** | * | * | 7 |
| Htun et al., 2021 | * | * |  | ** | ** | * | * | 8 |
| Islam et al., 2020 | * | * |  | ** | ** | * | * | 8 |
| Hu et al., 2021 |  | * |  | ** | ** | * | * | 7 |
| Kang et al., 2020 |  | * |  | ** | ** | * | * | 7 |
| Katz et al., 2020 |  |  |  | ** | ** | * | * | 6 |
| Ko et al., 2021 |  |  |  | ** | ** | * | * | 6 |
| Lai et al., 2020a |  | * | * | ** | ** | * | * | 8 |
| Lawson et al., 2021 |  |  |  | ** | ** | * | * | 6 |
| Le et al., 2022 |  |  |  | ** | ** | * | * | 6 |
| Liang et al., 2020 |  |  |  | ** | ** | * | * | 6 |
| S. Liu et al., 2021 |  | * |  | ** | ** | * | * | 7 |
| Lugito et al., 2021a |  |  |  | ** | ** | * | * | 6 |
| Ma et al., 2020 | * | * | * | ** | ** | * | * | 9 |
| Maehl et al., 2021 |  |  |  | ** | ** | * | * | 6 |
| Jiang et al., 2022 |  |  |  | ** | ** | * | * | 6 |
| Lu et al., 2022 |  | * |  | ** | ** | * | * | 7 |
| Mornioli et al., 2020 |  |  | * | ** | ** | * | * | 7 |
| Steward et al., 2021 | * | * | * | ** | * | * | * | 8 |
| Murphy et al., 2022b |  |  |  | ** | ** | * | * | 6 |
| Davis et al., 2021 |  |  |  | ** | ** | * | * | 6 |
| Chi et al., 2020 | * |  | * | ** | ** | ** | * | 9 |
| Chen and Li, 2021 | * | * | * | ** | ** | * | * | 9 |
| Cai et al., 2020 |  | * | * | * | ** | * | * | 7 |
| Buonsenso et al., 2022 |  |  |  | * | * | * | * | 4 |
| Bettinsoli et al., 2020 |  |  |  | ** | ** | * | * | 6 |
| Bella Nichole and Jonathan, 2020 |  | * | * | ** | ** | * | * | 8 |
| Angelina et al., 2021 |  |  |  | ** | * | * | * | 5 |
| Alshumrani et al., 2022 |  | * |  | ** | ** | * | * | 7 |
| AlAteeq et al., 2020 |  | * |  | ** | ** | * | * | 7 |
| He et al., 2022 | * | * | * | ** | ** | * | * | 9 |
| Gramaglia et al., 2022 | * | * | * | ** | ** | * | * | 9 |
| Gloster et al., 2020 |  | * |  | ** | ** | * | * | 7 |
| Gao et al., 2021 | * | * | * | ** | ** | * | * | 9 |
| Priyantini et al., 2021 |  |  |  | ** | ** | * | * | 6 |

Assigned 1 point (\*) or 2 points (\*\*) based on performance against a predetermined criterion on the scoring system

**Table 4:** Result of Quality Appraisal (Cohort Studies Quality Assessment: Max. Score = 9)

| Paper | Selection |  |  |  | Comparability | Outcome |  | Total score |
| --- | --- | --- | --- | --- | --- | --- | --- | --- |
|  | Representativeness | Non-exposed cohort | Ascertainment of exposure | Outcome present? | Confounding factors | Outcome assessment | Follow-up |  |
| Jakhar et al., 2021 | * | * | * |  | ** |  | * | 6 |
| Kalita et al., 2021 | * |  | * |  | ** |  | * | 5 |
| Severinsen et al., 2021 | * | * | * |  | ** |  | * | 6 |

Assigned 1 point (\*) or 2 points (\*\*) based on a met criterion on the scoring system

### Mental health assessment tools

In total, 62 mental health assessment tools were used. The most used tools include the Patient Health Questionnaire [PHQ] (41.7%), Generalized Anxiety Disorder Scale [GAD] (36%), 21-item Depression, Anxiety, and Stress [DASS-21] (13.9%), Impact of Event Scale [IES] (12.5%), Pittsburgh Sleep Quality Index [PSQI] (9.7%), Symptom Checklist [SCL] and the General Health Questionnaire [GHQ] (6.9% each). Three studies (4.1%) used a custom-made questionnaire that had the standard elements for the assessment of mental health conditions. The most studied mental health symptom was depression (73.6%) and anxiety (70.8%). Also, some assessed stress (41.6%), sleep issues/insomnia (26.4%), general mental health status (19.4%), general psychological states (13.8%), and post-traumatic stress disorder/symptoms (8.3%). Coping, fatigue, loneliness, and general well-being were also assessed. Specific tools used included PHQ-2/4/8/9 for depression, GAD-2/7 for anxiety, ISI for insomnia, IES/PSS for stress, CD-RISC resilience, PSQI for sleep quality, and DASS-21 as a stand-alone tool to measure anxiety, depression, and stress.

### Quality assessment

Using the NOS star rating as shown in Table 3, out of the 69 cross-sectional studies reviewed, 50 (72.5%) were of high quality, and the other 19 (27.5%) and the 3 cohort studies (Table **4**) reviewed were of moderate qualities (Fig 2).

**Fig 2:**
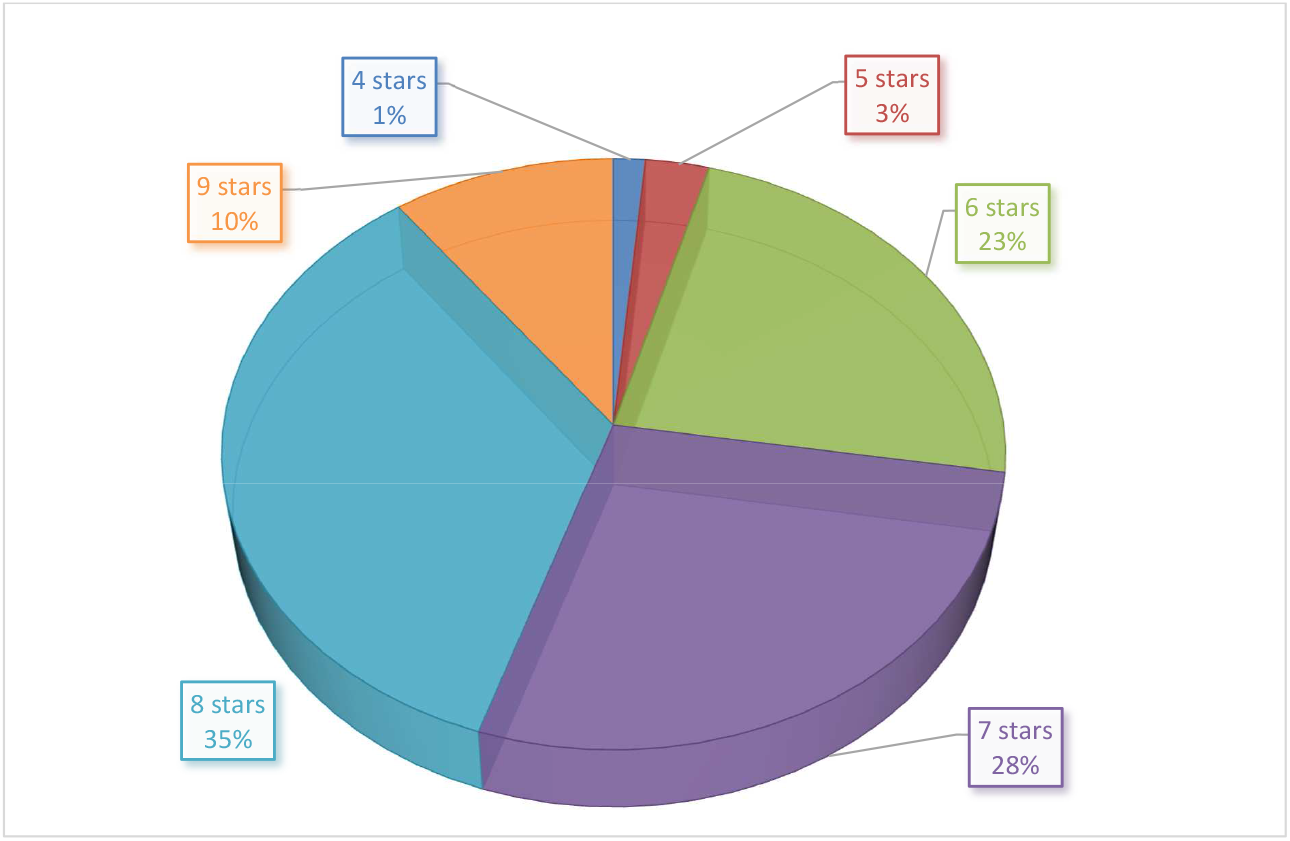
Pie chart showing the quality distribution of cross-sectional studies.

#### Prevalence Rate and Identified Risk Factors Mental Health Conditions

Table 5 represents the compiled risk factors for various health conditions across the studies. Various incident rates were also computed from different studies for mental health conditions, and the minimum, maximum, and arithmetic mean of prevalence was calculated for depression, anxiety, PTSD, sleep disorder, stress, psychological distress, and other studies which used general health conditions for showing the average pattern of prevalence.

**Table 5:** Prevalence rate and risk factors mental health conditions.

| Mental Health Condition | Incidence Rate (%) |  |  | Risk Factors |  |  |  |  |  |  |
| --- | --- | --- | --- | --- | --- | --- | --- | --- | --- | --- |
|  | Min | Max | Mean | Demographic-related | Relationship-related | Psychological-related | Occupation-related | Socioeconomic-related | Health-related | Information related |
| Depression | 5.00 | 76.48 | 34.75 | •Female gender (n=8)<br>•Male gender (n=4)<br>•Younger children (n=4)<br>•Older age (n=3)<br>•Divorcees (n=1)<br>•Proximity to high cases (n=5)<br>•Senior Students (n = 2)<br>•Urban Students (n = 1) | • Being quarantine (n = 1)<br>• Concern about family (n = 3)<br>• Link with someone infected/isolated (n = 1)<br>• Family history of hospitalization (n = 1)<br>• Living with a partner (n = 1)<br>• Lower family support (n = 1) | •Loneliness (n = 1)<br>•Pessimism (n = 1)<br>•Worry/fear (n = 6)<br>•High stress/distress (n = 1)<br>•Lower resilience (n = 1)<br>•Lower self-efficacy (n = 1) | •Shorter work experience (n =2)<br>•Working longer hours (n =2)<br>•Exposure to COVID-19 at work (n =1)<br>•Frontline workers (n =2)<br>•Nurses (n =1)<br>•Private sector workers (n =1) | •Losing job to pandemic (n =3)<br>•Financial concerns (n =1)<br>•Low social support (n =3)<br>•Lower income (n =2)<br>•Lower socioeconomic status (n =1)<br>•Living in a smaller home (n =2) | •Low health (n =1)<br>•History of mental disorders (n =3)<br>•Report of previous illness (n =1)<br>•Sleep issues (n =2)<br>•Presence of COVID-19 symptoms (n =1)<br>•Perceived exposure to COVID-19 (n =1) | •Lower knowledge of COVID-19 (n =2)<br>•More exposure to social media (n =3)<br>•Lower education (n =1)<br>•Higher education (n =1) |
| Anxiety | 5.63 | 80.5 | 35.63 | •Female gender (n =11)<br>•Male gender (n =1)<br>•Younger age (n =4)<br>•Older age (n =1)<br>•Divorcees (n =1)<br>•Senior students (n =2)<br>•Urban students (n =1) | •Concern for family members (n =2)<br>•Link with someone infected/isolated (n =1)<br>•Less family support (n =2)<br>•Family history of hospitalization (n =2) | •Depression (n =1)<br>•Helplessness (n =1)<br>•Loneliness (n =1)<br>•Worry/fear (n =4)<br>•High stress/distress (n =1)<br>•Lower resilience (n =1)<br>•Lower self-efficacy (n =1)<br>•Adverse childhood experiences (n =1) | •Frontline medical workers (n =3)<br>•Nurses (n =1)<br>•Lower work experience (n =1)<br>•Higher occupational exposure (n =1) | •Lower-income (n =2)<br>•Higher-income (n =1)<br>•Loss of job (n =2)<br>•Living in a smaller house (n =1)<br>•Lower socioeconomic status (n =1)<br>•Lower social support (n =2) | •Perceived exposure to COVID-19 (n =1)<br>•Being quarantined (n =1)<br>•History of hospitalization or previous illnesses (n =2)<br>•History of mental health conditions (n = 2)<br>•Presence of COVID-19 symptoms (n =1)<br>•Presence of underlying disease (n =1)<br>•Lower health (n =1)<br>•Lower sleep quality (n =3) | •Low education level (n =2)<br>•High education level (n =2)<br>•Low knowledge of COVID-19 (n =1)<br>•More exposure to social media (n =2) |
| PTSD | 9.1 | 65.0 | 37.06 | • Younger age (n =1)<br>• Middle age (n =1)<br>• Older age (n =1)<br>• Male gender (n =3)<br>• Female gender (n =2)<br>• Senior students (n =1)<br>• Urban students (n =1) | •Concern for family members (n =1)<br>•Link with infected/ isolated people (n =1)<br>•Less family support (n =1)<br>•More adverse childhood experiences (n =1) | •A higher number of stressors (n =1)<br>•Lower resilience (n =1)<br>•Lower self-efficacy (n =1) | •Less work experience (n =1)<br>•Working long hours (n =1)<br>•Nurses (n =1) | •Low social support (n =3) | •Presence of sleep issues (n =1)<br>•Perceived high risk of infection (n =1) |  |
| Sleep Disorder | 8.27 | 61.7 | 30.74 | •Male gender (n =2)<br>•Proximity to areas of high incidence (n =1) | •Concern about family members (n =1) | •Anxiety (n =1) | •Higher occupational exposure (n =1)<br>•Nurses (n =1)<br>•Healthcare assistants (n =1)<br>•Frontline medical workers (n =2)<br>•Workers with work experience ≥ 15 years (n =1) | •Losing job to pandemic (n =1) | •Being quarantined (n =1)<br>•Presence of underlying disease (n =1) |  |
| Stress | 4.9 | 70.1 | 34.88 | •Female gender (n =1)<br>•Younger age (n =2)<br>•Proximity to areas of high incidence (n =2)<br>•Early college years (1st/2nd) (n =1) | •Concern about family members (n =2)<br>•Lower family support (n =1) | •Fear/worry (n =1)<br>•Helplessness (n =1) | •Higher occupational exposure (n =1)<br>•Frontline workers (n =1) | •Lower socioeconomic status (n =1)<br>•Losing job to pandemic (n =1) | •Being quarantined (n =1)<br>•Previous illnesses/ medical history (n =2)<br>•Presence of COVID-19 symptoms (n =1)<br>•Perceived exposure to COVID-19 (n =1) | •More social media exposure (n =1)<br>•Low knowledge of COVID-19 (n =1) |
| Psychological Distress | 7 | 71.5 | 34.61 | •Female gender (n =3)<br>•Lower age (n =2)<br>•Proximity to areas of high incidence (n =1)<br>•Early college years (1st/2nd) (n =1) | •Lack of confidant (n =1)<br>•Frequent arguments with parents (n =1) | •Worry (n =2) | •Frontline healthcare workers (n =1) | •Low social support (n =1) | •Bad dietary patterns (n =1)<br>•Poor sleep quality (n =1)<br>•Presence of chronic disease (n =1)<br>•History of mental health conditions (n =2) |  |
| General Mental Health Conditions | 21.4 | 69.3 | 40.97 | •Female gender (n =8)<br>•Married (n =1)<br>•Divorced/ married (n =1)<br>•Single (n =1)<br>•Younger age (n =2)<br>•Older age (n =1)<br>•School closure/ lockdown (n =1)<br>•First=year students (n =1) | •Change of family habits as a result of the pandemic (n =1)<br>•Having to leave home frequently (n =1)<br>•Reduced family care (n =1)<br>•Reduced family visits to the hospital (n =1) | •Helplessness (n =1)<br>•Hopelessness (n =1)<br>•Concern for loved ones (n =1)<br>•Fear/worry (n =1)<br>•Severe stress levels (n =1)<br>•Perceived discrimination (n =1)<br>•Higher self-efficacy (n =1) | •Higher working experience (n =1)<br>•Working longer hours (n =1)<br>•Occupational stress (n =1)<br>•Working in healthcare (n =1)<br>•Nurse (n =2) | •Lower financial status (n =2)<br>•Lower-income range (n =2)<br>•Perceived lower social support (n =1)<br>•Less access to basic supplies (n =2)<br>•Loss of job to pandemic (n =1)<br>•Being employed (n =1) | •Report of poor health (n =1)<br>•Report of less sleep (n =1)<br>•Perceived exposure to COVID-19 (n =2)<br>•Testing negative for the virus (n =1) | •Increased exposure to health information (n =1)<br>•Lower coping mechanisms (n =2) |

#### Variations in the Incidence of Mental Health Conditions

Some epidemiological differences were observed in different population groups, places, and periods of study.

### Demographic Variations in Prevalence of Mental Health Conditions

Variations in the incidence of various mental health conditions among different demographic groups are depicted in Table 6.

**Table 6:**
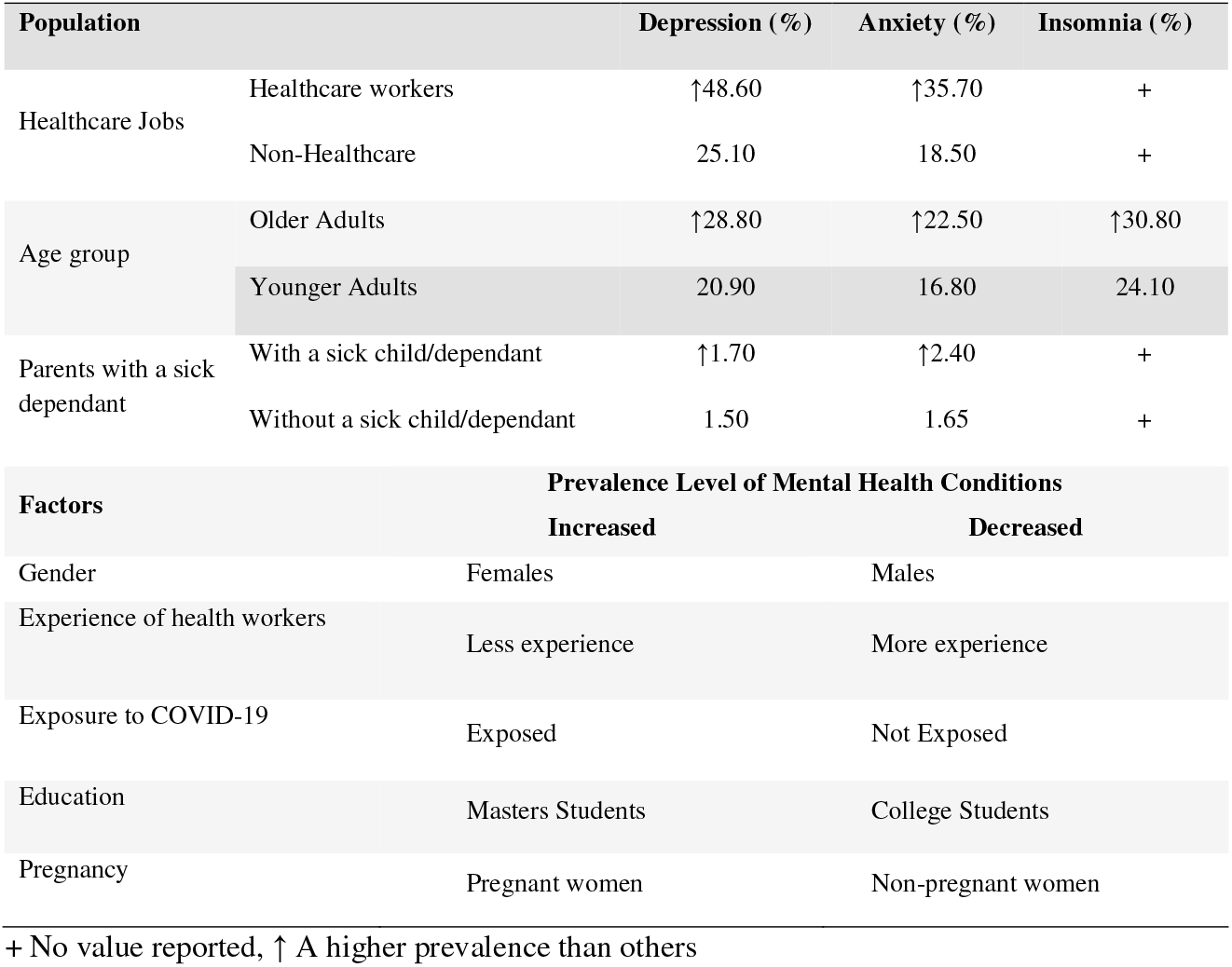
Variations of mental Health Conditions in Different Populations

### Variations in mental health prevalence over time

The prevalence of depression, anxiety, and PTSD were compiled for the studies conducted on them, which were mostly in 2020. Arithmetic means a calculation involving the groping of values over three months intervals, which was carried out on the reported prevalence from January to September 2020. Three months interval was selected because some months did not have any or enough studies conducted on the selected conditions. The resulting pattern showing the average dynamic of the prevalence is summarized in Fig. 3.

**Fig 3:**
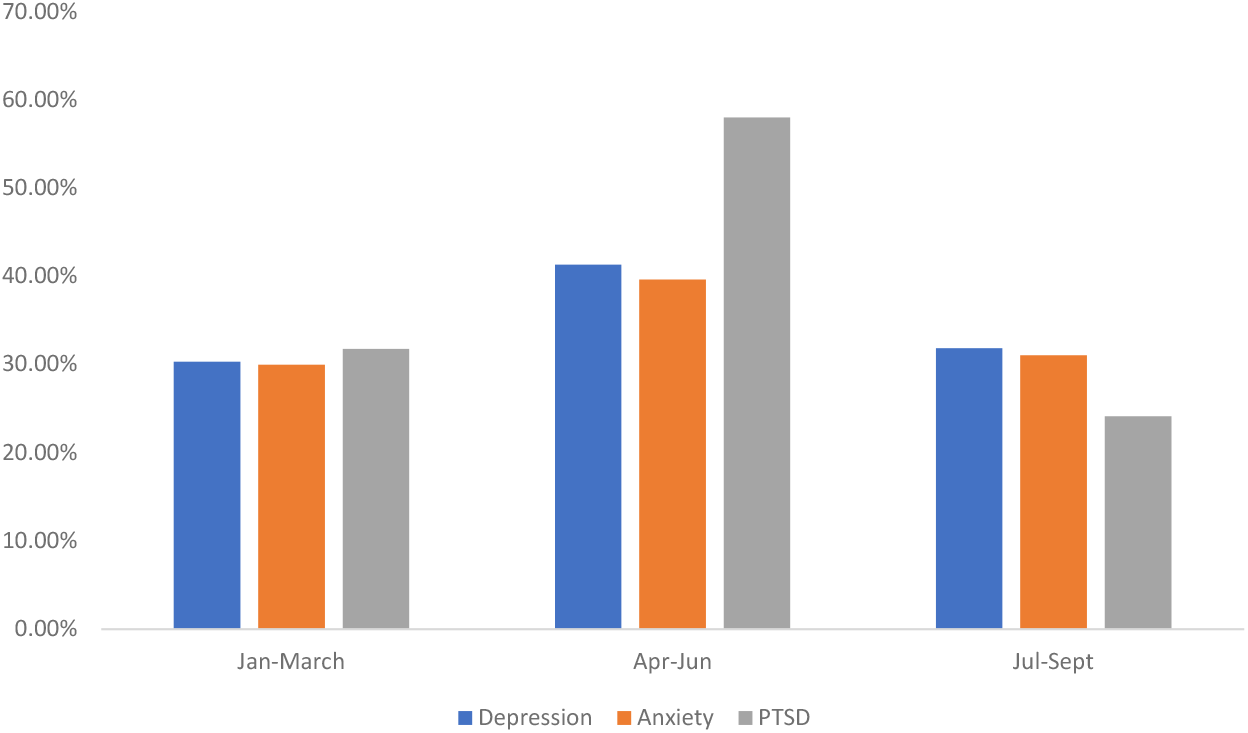
A Quarterly Representation of the Mean Prevalence of depression, anxiety, and PTSD from Jan. to Sept. 2020.

### Variations across countries and continents

Table 7 shows the maximum, mean, and minimum prevalence of mental health disorders across the assessed five continents. Only one study was found written on some disorders in some continents, while fewer than 5 were found in most other continents per disorder.

**Table 7:** Prevalence of Mental Health Challenges during the COVID-19 Pandemic in five Continents

| Continent | Psychological Distress | Stress | Sleep Disorder | PTSD | Anxiety | Depression | General Mental Health Conditions | Value |
| --- | --- | --- | --- | --- | --- | --- | --- | --- |
| Africa | 27.60 (n=1) | 28.60 (n=1) | - | - | 43.20(n=2) | 32.85(n=2) | 72.50*(n=1) | Mean |
|  | 27.60 | 28.60 | - | - | 52.00* | 46.30 | 72.50* | Max |
|  | 27.60+ | 28.60+ | - | - | 34.40 | 19.40 | 72.50+ | Min |
| Asia | 29.02(n=5) | 39.18(n=12) | 36.70(n=10) | 31.36(n=6) | 32.68(n=32) | 34.01(n=32) | 44.81(n=3) | Mean |
|  | 54.10* | 80.20* | 68.90* | 49.38 | 73.70* | 76.48* | 60.77* | Max |
|  | 7.00 | 6.40 | 8.27 | 9.10 | 5.63 | 5.00 | 27.05 | Min |
| Europe | - | 38.10(n=) | 16.00(n=1) | 53.17*(n=3) | 19.24(n=5) | 18.20(n=4) | 52.03*(n=3) | Mean |
|  | - | 38.10 | 16.00 | 65.00* | 42.00 | 28.90 | 70.30* | Max |
|  | - | 38.10 | 16.00 | 44.50 | 2.40 | 2.90 | 33.50 | Min |
| North America | 20.80(n=1) | - | - | - | 35.67*(n=3) | 49.55(n=3) | - | Mean |
|  | 20.80 | - | - | - | 69.10* | 77.50* | - | Max |
|  | 20.80+ | - | - | - | 41.40 | 7.90 | - | Min |
| South America | 62.80*(n=1) | - | - | - | 65.00(n=10) | 50.70(n=1) | - | Mean |
|  | 62.80 | - | - | - | 65.00 | 50.70 | - | Max |
|  | 62.80+ | - | - | - | 65.00+ | 50.70+ | - | Min |

+ Only one study was found here
\* Value is above 50% prevalence
- No value in cell

In selected eight (8) countries with the highest prevalence records during the pandemic, the chart below (Fig 4) contains the data on the prevalence of depression and anxiety

**Fig 4:**
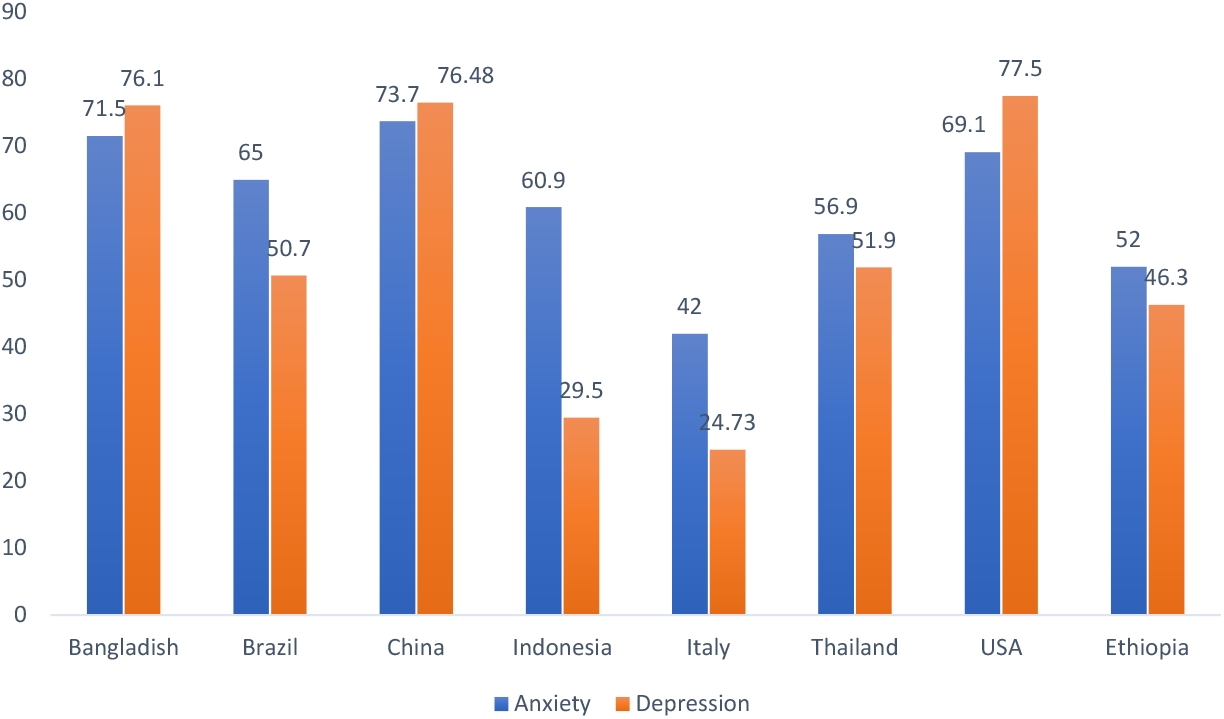
Highest prevalence recorded on mental health challenges during the COVID-19 Pandemic in eight selected countries.

## Discussion

The scoping review describes the prevalence of mental health disorders, the mental health tools used, and the risk factors identified by researchers during conditions during the COVID-19 pandemic.

### Mental health assessment tools

All reviewed studies used standard mental health assessment tools (89–93). The popular Patient Health Questionnaire (41.7%), and Generalized Anxiety Disorder Scale (36%) were the most applied tool by researchers in the assessment of mental health conditions. This strongly corresponds to the high number of studies that engaged in the assessment of depression, anxiety, and either stress or PTSD during the pandemic (23,25,28,40,42,94).

### Prevalence of mental disorders during the COVID-19 pandemic

Depression, anxiety, and stress were the most studied mental health conditions. Most studies reported a high (prevalence ≥ 50%) vulnerability to these three mental conditions (21, 22, 36, 39, 43, 44, 47, 57, 62, 74, 86) This finding is consistent with previous studies (95, 96).On the contrary, Jörns-Presentati et al., (94), reported a lower prevalence (29.0%) of depression. According to the WHO, the pandemic spiked with a 25% increase in the prevalence of mental disorders worldwide (7,95,96). Indeed, these differences were mostly attributed to the stress of COVID-19 by most of the studies. Some studies, however, reflected closer findings to Jörns-Presentati et al., (94), for mental health challenges with prevalence ranging between 20% and 49% for depression, anxiety, and stress. This included Alshumrani et al., (35), Bella Nichole and Jonathan, (33), Chi et al., (28), Naser et al., (65), Nguyen et al., (64), Simegn et al., (67), Zhang et al., (71), and various other studies. Most studies were conducted before the lockdown and during periods when the lockdown was being relaxed around the world. Some studies also reported a lower prevalence, especially those conducted in places or periods with lower cases of COVID-19 (54,58,75). Public crises can cause mental health disorders to rise way more than is naturally experienced among people, therefore, public health practitioners should be alert to the mental health of people and patients in crises.

### Factors associated with mental health disorder prevalence

This study revealed that many risk factors were associated with the presence of mental health mental disorders experienced during the COVID-19 pandemic. Most of the studies reported a higher prevalence of mental health challenges such as anxiety and depression among females who are usually more vulnerable to stress and psychological distress such as PTSD (73,97–99), in line with earlier studies (65,100–105). On the contrary Liu et al (44) and Oginni et al (106) found a higher prevalence of PTSD among males. Pregnant women and lactating mothers also showed a higher prevalence of mental disorders during the pandemic (107).

On the other hand, the higher prevalence of mental health conditions among females may also be attributable to the fact that higher numbers of the COVID-19 frontline workers such as nurses and other categories of caregivers are females who were faced with heightened COVID-19 challenges both at work and home during the pandemic (65,66,68,73,98,99,108,109). This finding is consistent with past studies (71,108,110) which found that health workers were vulnerable to the key risk factors for developing stress, anxiety, depression, and PTSD. Some studies reported that the length of one work experience, training, and support mechanisms helped reduce extreme burnout, psychological stress, and distress (111,112).

Equally noteworthy, people who shared proximity to places with higher COVID-19 cases had a higher level of mental health challenges (22,45,82). This finding is in line with the WHO observation that depression and anxiety disorders were higher in places with higher COVID-19 cases (113,114). This is supported by previous studies that showed that the fear and anxiety associated with the threatening numbers of cases and death in people’s neighborhood was seen earlier to increase the serious risk for mental breakdown (115) since a perceived or actual increase in the risk of exposure to COVID-19 was a major driver for adverse mental health (48,74).

Some studies reported that poor psychosocial support increased the prevalence and severity of mental health disorders among vulnerable population groups such as strictly isolated or heavily quarantined persons (116), individuals who lacked family support or care (117), elderly persons in need of nursing care (49,118), persons at risk of losing their jobs (33), persons without financial and social support (24,30,42,47,54,63,69,77,119), divorced persons (23,65,79), and relatives and guardians of sick persons (84). This finding indicates that social connections are a strong mechanism of survival and stress management for humans. Once this bond is disturbed by any stressors (120,121), such as done by COVID-19 pandemic, the mind is bound to be impacted if no interventions are put in place (8,117,122–125).

A few studies reported that living with a partner and being married were risk factors for mental health disorders, especially for individuals who also had mental or physical health challenges (71). During the Covid-19 pandemic, stressors from work and home were multi-factorial triggers to mental health distress (126,127), just as living with a partner with fears, mental stress, and emotional vulnerability, which was more prevalent during COVID-19, has been linked in the past to the presence of psychological stress (76,121,128).

Several articles found that one’s emotional or psychological state contributed to mental health conditions (52,119,129–133). Positive feelings such as hope, optimism, and self-efficacy were generally associated with better mental health status than helplessness, pessimism, worry/fear, distress, and anxiety (129,131,134,135). Ying Zhang et al. (73), however, reported that participants with higher self-efficacy had an increased risk of mental breakdown, similar to Khalil et al. (52,136), which explained that participants with higher self-efficacy reported having lower assertiveness, which was a stronger predictor of mental illnesses (134,135).

Similarly, one’s socioeconomic status was linked to an increased risk of mental health problems in almost all of the reviewed articles following similar findings (137–139). Sampaio et al., (98), reported a higher risk of depression among healthcare practitioners with higher incomes, which is justifiable looking at the fact that some healthcare workers doing overtime and extra shifts make more money, but at the risk of severe adverse health effects (98,139).

Persons with pre-existing mental and non-mental health issues, especially persons suffering from chronic illnesses, were more prone to higher mental health illnesses than otherwise healthy individuals (140–143), in agreement with the established relationships between health status and mental health (140) by studies such as MacMillan (144) and MHF (145). Mental health challenges among COVID-19 patients were higher than the general population in most of the studies (59,80,146,147), except Alshumrani et al., (35) who found that COVID-19 patients were less likely to suffer from mental health breakdown during the pandemic, which they attributed to factors such as lesser fear of unknown or increased confidence among COVID-19 survivors (35).

Although the impact of specific details of contents people got exposed to were not reported in any studies, some evidence showed that increased social media exposure was linked to an increase in the risk of mental health conditions (148–150). There was, however, no clear conclusion whether social media exposure led to an increase in mental health conditions. While some studies posited that social media can be a force for good when used properly, others suggested that the spread of uncensored content and unverified information would have been the reason behind the higher occurrence of mental disturbances among people with more social media usage (150–152).

### Trends in the prevalence of mental health conditions

Using the average values of the data collated the result showed that just as the pandemic grew stronger, the global prevalence of mental health conditions rose sharply from 30.31%, 29.97%, and 31.74% to 41.31%, 39.61%, and 58% for depression, anxiety, and post-traumatic stress disorder respectively (7,95,96,113,147). A decline to 31.83%, 31.03%, and 24.10% was also observed for the three disorders as the cases and restrictions started reducing in various places (33,42,73,87,95,96,113), although the impact continued to linger (95). There were variations in mental health prevalence across different population groups from 24 different countries in Asia, Africa, North America, South America, and Europe (Table 7). The differences observed in countries were mostly related to outbreak severity, degree of government-imposed restrictions, and socioeconomic status of the region (22,60,61,66,85,86).

### Variations due to sociodemographic attributes

The females (65), frontline workers (109,110), people who were ill (41,66), who work long hours (68,153), whose job increased their exposure to COVID-19 (48,74), were living in proximity to COVID-19 cases, (22,45,82), were young or older (4,5,35,62,83,84,107,116,118,154–159), had lower economic & education status, weak psychological makeup, and low social supports (142) had higher levels of mental health conditions during the pandemic (7,113,148), which serves as a call to set up protective measures for this population during any interventions.

### Geographical variations

Table 7 shows that the highest prevalence of depression and anxiety was in North America, precisely the United States (47), followed by China (39), and Bangladesh (21). Stress and sleep disorders were highest in Asia, especially Thailand (57) and Bangladesh (21) respectively. Also, the prevalence of psychological distress was highest in Brazil (69) which represented South America, while general mental health disorders or conditions showed up more in Africa (87), followed closely by Europe (13,56) (Fig 4). The review found that at the country level, Bangladesh, the USA, and China were at the top of the list for both depression and anxiety during the pandemic (21,43,119).

### Time-related variations of mental health challenges

In Bangladesh Islam et al., (21), found a significantly higher level of mental health than those conducted before and after the peak of COVID-19 incidence. In general, the reported prevalence of most mental disorders rose steadily with the advent of the pandemic until the middle of the year 2020. This coincides with periods with higher COVID-19 cases and deaths with stricter lockdown measures (160) (Fig 4). After this peak period, a steady decline was observed in the last 3 months, with values falling below those obtained during the onset of the pandemic. This decline followed the period the governments began to ease off the COVID-19 restrictions (2,13,95,96,160–162). Although there is a wide view that the mental health prevalence naturally reduced as COVID-19 eased off or people adjusted to the new reality, it is important to bear in mind that the observed prevalence after the strict lockdown was still higher than before (80,95).

### Gaps in literature

Although COVID-19 and mental health are crucial global issues, most studies were conducted in Asia, with few from Europe, while North & South America and Africa had a very limited number. Based on the inclusion criteria for this review, no relevant articles were found in Australia or Antarctica. This partially limited the ability to draw a clear line on the global prevalence of mental health conditions.

Few studies investigated the impacts of COVID-19 on the mental health of other vulnerable populations, such as students, pregnant women, children, the elderly, and persons with chronic diseases. Few studies examined the specific relationship between government restrictions and mental health conditions. More research is needed to examine these issues in detail to guide future interventions by governments and policymakers.

### Conclusion and recommendations

#### Conclusion

This review found that various standard tools were used to assess mental health disorders during the COVID-19 pandemic. These included the Patient Health Questionnaire, Generalized Anxiety Disorder Scale, 21-item Depression, Anxiety, and Stress, Impact of Event Scale, and Pittsburgh Sleep Quality Index. The prevalence of mental health conditions increased during the COVID-19 pandemic and decreased as the COVID-19 prevalence reduced. Also, the relaxation of the COVID-19 restrictions contributed to a decrease in the prevalence rate of mental health conditions. The review showed that one’s profession, occupation, gender, age, marital status, family relationships, socioeconomic status, access to information, psychological makeup, and longstanding health status, played important parts in the development of mental health conditions during the pandemic. Healthcare workers were more prone to the challenges as they were highly strained and faced by the pandemic than many other professions.

Although the number of articles from each varied, there were observed differences in the reported prevalence of mental health conditions from each continent and between countries assessed. It is clear, therefore, that the pandemic caused a significant rise in mental health challenges across the world which requires critical attention for better local and global health policies and processes management.

### Recommendations

- Governments and policymakers in public and private organizations should increase social protection as it is an essential ingredient in helping the public cope with such critical events.
- Efforts should be made towards putting mechanisms in place to mitigate mental health challenges during public health interventions in the future.
- More attention should be paid to providing support and training on how to improve their coping mechanisms during public health challenges.
- Further studies should investigate the effectiveness of interventions for reducing the prevalence and risk factors for mental health conditions in a public health crisis.
- More studies should continue to focus on the trends in mental health conditions, because the COVID-19 disease may have reduced, but the health impacts might linger
- While more studies on health crises relative to public health outbreaks and interventions are needed in Africa, America (North and South), and even Europe on mental, Australia and Antarctica need to be researched or reviewed for similar circumstances.
- Areas such as economic and social well-being, and stress-related illnesses such as hypertension, metabolic disorders, and gastrointestinal diseases which are possibly impacted by the pandemic should be further investigated or reviewed.

## Data Availability

This article is a scoping review. All sources reviewed and cited are listed in the reference list of this work. The methods section also has the steps followed in obtaining and screening the articles for review.

## Limitations

The review considered articles written in the English language, which may limit the generalizability of the study findings to the non-English speaking regions. The timeframe was also from March 2020 to July 2022, hence all other studies before and after this period will have findings that may support or contradict this study. The review included only open-source articles and did not include any articles that required payments or prior consent before review. This may have also limited the content of this review and the generalizability of the findings.

## Abbreviations

ACE: Adverse Childhood Experience
ADQ: Author Designed Questionnaire
AIS: Athens Insomnia Scale
ASDS: Acute Stress Disorder
AUDIT: Alcohol Use Disorders Identification Test
BAI: Beck Anxiety Inventory
BDI-II: Beck Depression Inventory-II
Brief COPE: Brief Coping Orientation to Problem Experienced
BRCS: Brief Resilience Coping Scale
CDI-S: Children’s Depression Inventory-Short Form
CCMH: Copenhagen Corona-Related Mental Health Questionnaire
CES-D: Center for Epidemiologic Studies Depression scale
CMD: Common Mental Health Diseases
CMHDQA-4: Four-item Common Mental Health Disorder Questionnaire Anxiety subscale
CSES: Coping Self-Efficacy Scale
CD-RISC: Connor-Davidson Resilience Scale
CD-RISK-10: Abbreviated Version of the Connor–Davidson Resilience Scale
CDI: Child Depression Inventory
COVID-19: Coronavirus Disease 2019
DAR-5: Dimensions of Anger Reactions-Revised
DASS-21: 21-item Depression, Anxiety, and Stress Scale
FSS: Fatigue Severity Scale
GAD-7: 7-item Generalized Anxiety Disorder scale
GHQ-12: General Health Questionnaire-12
GPS: Global Psychotrauma Screen
HADS: Hospital Anxiety and Depression Scale
IES: Impact of Event Scale
IES-R: 22-item Impact of Event Scale-Revised
ISI: Insomnia Severity Index
K-10: Kessler Psychological Distress Scale
LCKRS-2: Long COVID Kids Rapid Survey 2
MBI: Maslach Burnout Inventory
MHC-SF: Mental Health Continuum Short Form
MSBS: Multidimensional State Boredom Scale
NPI-Q: Neuropsychiatric Inventory Questionnaire
PANAS: Positive and Negative Affect Scale
PCL: Abbreviated PTSC Checklist
PCL-5: PTSD Checklist
PC-PTSD-5: Posttraumatic Stress Symptoms scale
PCQ: Psychological Capital Questionnaire
PHQ-4: 4-item Patient Health Questionnaire
PHQ-9: 9-point Patient Health Questionnaire
PROMIS: Patient-Reported Outcomes Measurement Information System
PSC: Pediatric Symptom Checklist
PSQI: Pittsburgh Sleeping Quality Index
PSS: Perceived Stress Level
PSS-10: 10-item Perceived Stress Scale
PSQI: Pittsburgh Sleep Quality Index
PSWQ-C: Penn State Worry Questionnaire for Children
PTGI: Post Traumatic Growth Inventory
SAS: Self-rating Anxiety Scale
SCARED: Screen for Child Anxiety-Related Emotional Disorders
SCL-90: Symptom checklist 90
SCSQ: Short Coping Style Questionnaire
SDS: Self-rating Depression Scale
SRSS: Self-Rating Scale of Sleep
SRQ-20: 20-item Self Reporting Questionnaire
STAI-Y: State-Trait Anxiety Inventory-Form Y
STATE-A: Anxiety subsequent to a specific situation
ULS-3: 3-item UCLA Loneliness Scale
ULS-8: 8-item UCLA Loneliness Scale
VTQ: Vicarious Traumatization Questionnaire
Z-SAS: Zung Self-Rating Anxiety Scale

## Acknowledgments

The first author would like to extend his sincere appreciation to the management and staff of Liverpool John Moores University and UNICAF University for their scholarship support towards his postgraduate studies and to his co-author, Dr. Frances Ncube, for his relentless guidance and contributions. The authors wish to thank Ms. Brontie A. Duncan for proofreading this article. The support and encouragement from Mrs. Felicia Otoboyor, Mr. Ndidi L. Otoboyor, Mr. Oghosa G. Josiah, Miss. Chinelo Uzor, Ms. Doren Francis, and Mr. Joshua Okonkwo are much appreciated.

## References

1. WHO. WHO Coronavirus Disease (COVID-19) Dashboard With Vaccination Data | WHO Coronavirus (COVID-19) Dashboard With Vaccination Data [Internet]. World Health Organization. 2021. p. 1–5. Available from: https://covid19.who.int/

2. IMF. Policy Responses to COVID19 [Internet]. International Monetary Fund Policy Tracker. 2021. Available from: https://www.imf.org/en/Topics/imf-and-covid19/Policy-Responses-to-COVID-19

3. Kupferschmidt K, Cohen J. China’s aggressive measures have slowed the coronavirus. They may not work in other countries. Science. 2020;

4. Courtney D, Watson P, Battaglia M, Mulsant BH, Szatmari P. COVID-19 Impacts on Child and Youth Anxiety and Depression: Challenges and Opportunities. Can J Psychiatry. 2020;65(10):688–91.

5. Shah S, Kaul A, Shah R, Maddipoti S. Impact of Coronavirus Disease 2019 Pandemic and Lockdown on Mental Health Symptoms in Children. Indian Pediatr. 2021;58(1):75–6.

6. UN. COVID-19 and the Need for Action on Mental Health 13 M AY 2 0 2 0. United Nations Policy Brief. 2020.

7. WHO. COVID-19 pandemic triggers 25% increase in prevalence of anxiety and depression worldwide [Internet]. World Health Organization News Release. 2022. Available from: https://www.who.int/news/item/02-03-2022-covid-19-pandemic-triggers-25-increase-in-prevalence-of-anxiety-and-depression-worldwide

8. Xiong J, Lipsitz O, Nasri F, Lui LMW, Gill H, Phan L, et al. Impact of COVID-19 pandemic on mental health in the general population: A systematic review. J Affect Disord. 2020;277:55–64.

9. Hannemann J, Abdalrahman A, Erim Y, Morawa E, Jerg-Bretzke L, Beschoner P, et al. The impact of the COVID-19 pandemic on the mental health of medical staff considering the interplay of pandemic burden and psychosocial resourcesA rapid systematic review. Doering S, editor. PLOS ONE. 2022;17(2):e0264290–e0264290.

10. Murphy D, Williamson C, Baumann J, Busuttil W, Fear NT. Exploring the impact of COVID-19 and restrictions to daily living as a result of social distancing within veterans with pre-existing mental health difficulties. BMJ Mil Health. 2022;168(1):29–33.

11. Jones K, Mallon S, Schnitzler K. A Scoping Review of the Psychological and Emotional Impact of the COVID-19 Pandemic on Children and Young People: 2021;1–25.

12. Panchal N, Kamal R, Orgera K. The implications of COVID-19 for mental health and substance use. Kais Fam Found. 2020;21.

13. ONS. Coronavirus and anxiety, Great Britain - Office for National Statistics. Office for National Statistics. 2020.

14. Tricco AC, Lillie E, Zarin W O KK, Colquhoun H, Levac D, et al. PRISMA Extension for Scoping Reviews (PRISMA-ScR): Checklist and Explanation. Ann Intern Med. 2018;169(7):467–73.

15. Peters M, Godfrey C, McInerney P, Munn Z, Trico A, Khalil H. Chapter 11: Scoping Reviews BT - JBI Manual for Evidence Synthesis. In: JBI Manual for Evidence Synthesis. 2020.

16. Arksey H, O’Malley L. Scoping studies: towards a methodological framework. Int J Soc Res Methodol. 2005;8(1):19–32.

17. Naidoo K, van Wyk J. Protocol for a scoping review of age-related health conditions among geriatric populations in sub-Saharan Africa. Syst Rev. 2019;8(1).

18. Negera Getandale. Re: Can someone explain to me how can I use the Newcastle-Ottawa Scale(NOS) for assessing the Quality? [Internet]. 2020. Available from: https://www.researchgate.net/post/Can_someone_explain_to_me_how_can_I_use_the_Newcastle-Ottawa_ScaleNOS_for_assessing_the_Quality/5ec62f0864c32b284a447d0f/citation/download

19. Modesti PA, Reboldi G, Cappuccio FP, Agyemang C, Remuzzi G, Rapi S, et al. Panethnic Differences in Blood Pressure in Europe: A Systematic Review and Meta-Analysis. PLOS ONE. 2016;11(1):e0147601–e0147601.

20. Penson DF, Krishnaswami S, Jules A, Seroogy JC, McPheeters ML. Evaluation and Treatment of Cryptorchidism [Internet]. Rockville (MD): Agency for Healthcare Research and Quality (US); 2012 [cited 2022 Nov 19]. (AHRQ Comparative Effectiveness Reviews). Available from: http://www.ncbi.nlm.nih.gov/books/NBK115847/

21. Islam MS, Sujan MSH, Tasnim R, Sikder MT, Potenza MN, van Os J. Psychological responses during the COVID-19 outbreak among university students in Bangladesh. PLoS One. 2020;15(12):e0245083–e0245083.

22. He Q, Fan B, Xie B, Liao Y, Han X, Chen Y, et al. Mental health conditions among the general population, healthcare workers and quarantined population during the coronavirus disease 2019 (COVID-19) pandemic. Psychol Health Med. 2022;27(1):186–98.

23. Gramaglia C, Bazzano S, Gambaro E, Cena T, Azzolina D, Costa A, et al. Mental Health Impact and Burnout in Critical Care Staff During Coronavirus Disease 2019 Outbreak. Turk J Anaesthesiol Reanim. 2022;50(Supp1):S34-S41-S34–41.

24. Gloster AT, Lamnisos D, Lubenko J, Presti G, Squatrito V, Constantinou M, et al. Impact of COVID-19 pandemic on mental health: An international study. Francis JM, editor. 2020;15(12):e0244809–e0244809.

25. Gao J, Wang F, Guo S, Hu F. Mental Health of Nursing Students amid Coronavirus Disease 2019 Pandemic. Front Psychol. 2021;12:699558.

26. Priyantini D, Nursalam N, Sukartini T, Diah P, Nursalam N, Tintin S, et al. Analysis of Factors Affecting the Mental Health Crisis of Coronavirus Disease Infection in Java Island. J Ners. 2021;16(1):60–6.

27. Davis EJ, Amorim G, Dahn B, Moon TD. Perceived ability to comply with national COVID-19 mitigation strategies and their impact on household finances, food security, and mental well-being of medical and pharmacy students in Liberia. Atiqul Haq SM, editor. PLOS ONE. 2021;16(7):e0254446–e0254446.

28. Chi X, Becker B, Yu Q, Willeit P, Jiao C, Huang L, et al. Prevalence and Psychosocial Correlates of Mental Health Outcomes Among Chinese College Students During the Coronavirus Disease (COVID-19) Pandemic. Front Psychiatry. 2020;11:803.

29. Chen Y, Li W. Influencing Factors Associated With Mental Health Outcomes Among Dental Medical Staff in Emergency Exposed to Coronavirus Disease 2019: A Multicenter Cross-Sectional Study in China. Front Psychiatry. 2021;12:736172.

30. Cai W, Lian B, Song X, Hou T, Deng G, Li H. A cross-sectional study on mental health among health care workers during the outbreak of Corona Virus Disease 2019. Asian J Psychiatry. 2020;51:102111.

31. Buonsenso D, Pujol FE, Munblit D, Pata D, McFarland S, Simpson FK. Clinical characteristics, activity levels and mental health problems in children with long coronavirus disease: a survey of 510 children. Future Microbiol [Internet]. 2022; Available from: http://www.epistemonikos.org/documents/86a3e7bf73e85c2cbc380c61272206633634e941

32. Bettinsoli ML, Di Riso D, Napier JL, Moretti L, Bettinsoli P, Delmedico M, et al. Mental Health Conditions of Italian Healthcare Professionals during the COVID-19 Disease Outbreak. Appl Psychol Health Well-Being. 2020;12(4):1054–73.

33. Bella Nichole K, Jonathan K. Mental health outcomes and associations during the coronavirus disease 2019 pandemic: A cross-sectional survey of the US general population. medRxiv [Internet]. 2020; Available from: http://www.epistemonikos.org/documents/db89f399cfe5d9bfda8a80624fb94f7afba8e7ed

34. Angelina S, Kurniawan A, Agung FH, Halim DA, Wijovi F, Jodhinata C, et al. Adolescents’ mental health status and influential factors amid the Coronavirus Disease pandemic. Clin Epidemiol Glob Health. 2021;12:100903.

35. Alshumrani R, Qanash S, Aldobyany A, Alhejaili F, AlQassas I, Shabrawishi M, et al. Sleep quality and mental health in coronavirus disease 2019 patients and general population during the pandemic. Ann Thorac Med. 2022;17(1):21–7.

36. AlAteeq DA, Aljhani S, Althiyabi I, Majzoub S. Mental health among healthcare providers during coronavirus disease (COVID-19) outbreak in Saudi Arabia. J Infect Public Health [Internet]. 2020;13(10). Available from: http://www.epistemonikos.org/documents/4d867f54d8a5200ab841d0236414f24f0e8c4798

37. Steward M, Moses M, Chiluba M, Zikria S, Aubrey Chichoni K, Derick M, et al. Impact of the Coronavirus Disease (COVID-19) on the Mental Health and Physical Activity of Pharmacy Students at the University of Zambia: A Cross-Sectional Study. medRxiv [Internet]. 2021; Available from: http://www.epistemonikos.org/documents/f7512d15985efaf1dc7e307d97bd8dfc33c19c4c

38. Morniroli D, Consales A, Colombo L, Bezze E, Zanotta L, Plevani L, et al. Newly Mothers’ Mental Health and Breastfeeding Rates During 2019 Coronavirus Disease Outbreak. ResearchSquare [Internet]. 2020; Available from: http://www.epistemonikos.org/documents/72c75f4c3b03aada50aea0cd601d591d483474bb

39. Lu T, Yu Y, Zhao Z, Guo R. Mental Health and Related Factors of Adolescent Students During Coronavirus Disease 2019 (COVID-19) Pandemic. Psychiatry Investig [Internet]. 2022; Available from: http://www.epistemonikos.org/documents/0a1f02e8667890d55649f1ce762a4d6ff95c893f

40. Jiang YF, Chen JQ, Wang YG, Zhang XD, Hong WK. Sleep quality and mental health status of healthcare professionals during the outbreak of coronavirus disease 2019 (COVID-19). Psychol Health Med. 2022;27(2):488–95.

41. Jakhar J, Biswas PS, Kapoor M, Panghal A, Meena A, Fani H, et al. Comparative study of the mental health impact of the COVID-19 pandemic on health care professionals in India. Future Microbiol. 2021;16(16):1267–76.

42. Ma Z, Zhao J, Li Y, Chen D, Wang T, Zhang Z, et al. Mental health problems and correlates among 746 217 college students during the coronavirus disease 2019 outbreak in China. Epidemiol Psychiatr Sci. 2020;29:e181–e181.

43. Lugito NPH, Damay V, Chyntya H, Sugianto N. Social media exposure and mental health problems during coronavirus disease 2019 pandemic in Indonesia. J Educ Health Promot. 2021;10(1):200.

44. Liu S, Yang L, Zhang C, Xu Y, Cai L, Ma S, et al. Gender differences in mental health problems of healthcare workers during the coronavirus disease 2019 outbreak. J Psychiatr Res. 2021;137:393–400.

45. Liang Y, Wu K, Zhou YY, Huang X, Zhou YY, Liu Z. Mental Health in Frontline Medical Workers during the 2019 Novel Coronavirus Disease Epidemic in China: A Comparison with the General Population. 2020;17(18). Available from: http://www.epistemonikos.org/documents/fd803fb1d260c02351a74d5fd9e85c4c1a7a2bdc

46. Le PD, Misra S, Hagen D, Wang SM, Li T, Brenneke SG, et al. Coronavirus disease (COVID-19) related discrimination and mental health in five U.S. Southern cities. Stigma Health [Internet]. 2022; Available from: http://doi.apa.org/getdoi.cfm?doi=10.1037/sah0000351

47. Lawson AK, McQueen DB, Swanson AC, Confino R, Feinberg EC, Pavone ME. Psychological distress and postponed fertility care during the COVID-19 pandemic. J Assist Reprod Genet. 2021;38(2):333–41.

48. Lai J, Ma S, Wang Y, Cai Z, Hu J, Wei N, et al. Factors associated with mental health outcomes among health care workers exposed to coronavirus disease 2019. JAMA Netw Open. 2020;3(3):e203976–e203976.

49. Ko M, Cho HM, Park J, Chi S, Han C, Yi HS, et al. Impact of the Coronavirus Disease Pandemic on Mental Health among Local Residents in Korea: a Cross Sectional Study. J Korean Med Sci. 2021;36(46):e322–e322.

50. Katz P, Pedro S, Wipfler K, Simon T, Shaw Y, Cornish A, et al. Changes in Mental Health During the COVID-19 Pandemic Among Individuals with Rheumatic Disease [abstract]. Arthritis Rheumatol. 2020;10:72.

51. Kalita J, Tripathi A, Dongre N, Misra UK. Impact of COVID-19 pandemic and lockdown in a cohort of myasthenia gravis patients in India. Clin Neurol Neurosurg. 2021;202:106488.

52. Hu Q, Hu X, Zheng B, Li L. Mental Health Outcomes Among Civil Servants Aiding in Coronavirus Disease 2019 Control. Front Public Health. 2021;9:601791.

53. Htun YM, Thiha K, Aung A, Aung NM, Oo TW, Win PS, et al. Assessment of depressive symptoms in patients with COVID-19 during the second wave of epidemic in Myanmar: A cross-sectional single-center study. Mitra P, editor. PLOS ONE. 2021;16(6):e0252189– e0252189.

54. Hou T, Zhang T, Cai W, Song X, Chen A, Deng G, et al. Social support and mental health among health care workers during Coronavirus Disease 2019 outbreak: A moderated mediation model. PloS One. 2020;15(5):e0233831–e0233831.

55. Shen H, Wang H, Zhou F, Chen J, Deng L, Haiyan S, et al. Mental health status of medical staff in the epidemic period of coronavirus disease 2019. Zhong Nan Da Xue Xue Bao Yi Xue Ban. 2020;45(6):633–40.

56. Severinsen ER, Kähler LKA, Thomassen SE, Varga TV, Fich Olsen L, Hviid KVR, et al. Mental health indicators in pregnant women compared with women in the general population during the coronavirus disease 2019 pandemic in Denmark. Acta Obstet Gynecol Scand. 2021;100(11):2009–18.

57. Ruengorn C, Awiphan R, Wongpakaran N, Wongpakaran T, Nochaiwong S, Outcomes H, et al. Association of job loss, income loss, and financial burden with adverse mental health outcomes during coronavirus disease 2019 pandemic in Thailand: A nationwide cross-sectional study. Depress Anxiety [Internet]. 2021;38(6). Available from: http://www.epistemonikos.org/documents/11d6a247e4d3c5150b6315a5f8eddd0f9c6b9bed

58. Pieh C, Budimir S, Probst T. The effect of age, gender, income, work, and physical activity on mental health during coronavirus disease (COVID-19) lockdown in Austria. J Psychosom Res. 2020;136:110186.

59. Qin X, Shu K, Wang M, Chen W, Huang M, Yang A, et al. Mental health status of patients with coronavirus disease 2019 in Changsha. Zhong Nan Da Xue Xue Bao Yi Xue Ban. 2020;45(6):657–64.

60. Rossi R, Socci V, Pacitti F, Di Lorenzo G, Di Marco A, Siracusano A, et al. Mental Health Outcomes Among Frontline and Second-Line Health Care Workers During the Coronavirus Disease 2019 (COVID-19) Pandemic in Italy. JAMA Netw Open. 2020;3(5):e2010185– e2010185.

61. Prati G. Mental health and its psychosocial predictors during national quarantine in Italy against the coronavirus disease 2019 (COVID-19). Anxiety Stress Coping. 2021;34(2):145– 56.

62. Penteado CT, Loureiro JC, Pais MV, Carvalho CL, Sant’Ana LFG, Valiengo LCL, et al. Mental Health Status of Psychogeriatric Patients During the 2019 New Coronavirus Disease (COVID-19) Pandemic and Effects on Caregiver Burden. Front Psychiatry. 2020;11:578672.

63. Norhayati MN, Che Yusof R, Azman MY. Vicarious traumatization in healthcare providers in response to COVID-19 pandemic in Kelantan, Malaysia. Brownie SM, editor. PLOS ONE. 2021;16(6):e0252603–e0252603.

64. Nguyen PTL, Nguyen TBL, Pham AG, Duong KNC, Gloria MAJ, Vo TV, et al. Psychological Stress Risk Factors, Concerns and Mental Health Support Among Health Care Workers in Vietnam During the Coronavirus Disease 2019 (COVID-19) Outbreak. Front Public Health. 2021;9:628341.

65. Naser AY, Dahmash EZ, Al-Rousan R, Alwafi H, Alrawashdeh HM, Ghoul I, et al. Mental health status of the general population, healthcare professionals, and university students during 2019 coronavirus disease outbreak in Jordan: A cross-sectional study. Brain Behav. 2020;10(8):e01730–e01730.

66. Shi L, Lu ZA, Que JY, Huang XL, Liu L, Ran MS, et al. Prevalence of and Risk Factors Associated With Mental Health Symptoms Among the General Population in China During the Coronavirus Disease 2019 Pandemic. JAMA Netw Open. 2020;3(7):e2014053– e2014053.

67. Simegn W, Dagnew B, Yeshaw Y, Yitayih S, Woldegerima B, Dagne H. Depression, anxiety, stress and their associated factors among Ethiopian University students during an early stage of COVID-19 pandemic: An online-based cross-sectional survey. PLoS One. 2021;16(5):e0251670–e0251670.

68. Xingyue S, Wenning F, Xiaoran L, Zhiqian L, Rixing W, Ning Z, et al. Mental Health Status of Medical Staff in Emergency Departments During the Coronavirus Disease 2019 Epidemic in China. SSRN [Internet]. 2020; Available from: http://www.epistemonikos.org/documents/5d65132e88deb3523415a99cc1c253b9e428eca3

69. Teixeira L de AC, Costa RA, de Mattos RMPR, Pimentel D. Brazilian medical students mental health during coronavirus disease 2019 pandemic. J Bras Psiquiatr. 2021;70(1):21– 9.

70. Zheng Y, Wang L, Feng L, Ye L, Zhang A, Fan R. Sleep quality and mental health of medical workers during the coronavirus disease 2019 pandemic. Sleep Biol Rhythms. 2021;19(2):173–80.

71. Zhang X, Zhao K, Zhang G, Feng R, Chen J, Xu D, et al. Occupational Stress and Mental Health: A Comparison Between Frontline Medical Staff and Non-frontline Medical Staff During the 2019 Novel Coronavirus Disease Outbreak. Front Psychiatry. 2020;11:555703.

72. Zhang GY, Shen L, Liu Q, Lin JY, Si TM, Yan L. Mental health outcomes among patients from Fangcang shelter hospitals exposed to coronavirus disease 2019: An observational cross-sectional study. Chronic Transl Med. 2021;7(1):57–64.

73. Zhang Y, Tian L, Li W, Wen X, Wu H, Gong R, et al. Mental health status among Chinese healthcare-associated infection control professionals during the outbreak of coronavirus disease 2019: A national cross-sectional survey. Medicine (Baltimore). 2021;100(5):e24503–e24503.

74. Zhang Y, Li D, Ouyang X, Bai H, Zhao L, Shi Y, et al. Mental Health Differences in Healthcare Workers Exposed to Different Risks During the Coronavirus Disease 2019 Pandemic. Front Psychiatry. 2022;13:827076.

75. Zhan J, Sun S, Xie L, Wen Y, Fu J. Medical students’ mental health, professional pride, and intention to work in the front-line during coronavirus disease 2019 pandemic. Zhong Nan Da Xue Xue Bao Yi Xue Ban. 2020;45(6):649–56.

76. Ying Y, Ruan L, Kong F, Zhu B, Ji Y, Lou Z. Mental health status among family members of health care workers in Ningbo, China, during the coronavirus disease 2019 (COVID-19) outbreak: a cross-sectional study. BMC Psychiatry. 2020;20(1):379.

77. Xiaoxv Y, Jing W, Jie F, Zhenyuan C, Nan J, Jianxiong W, et al. The Impact of the Corona Virus Disease 2019 Outbreak on Chinese Residents’ Mental Health. SSRN Electron J [Internet]. 2020; Available from: http://www.epistemonikos.org/documents/39309f944d6088fa901a40845c99c9fa0e956cbb

78. Yang KH, Wang L, Liu H, Li LX, Jiang XL. Impact of coronavirus disease 2019 on the mental health of university students in Sichuan Province, China: An online cross-sectional study. Int J Ment Health Nurs. 2021;30(4):875–84.

79. Xu Z, Zhang D, Xu D, Li X, Xie YJ, Sun W, et al. Loneliness, depression, anxiety, and post-traumatic stress disorder among Chinese adults during COVID-19: A cross-sectional online survey. PLoS One. 2021;16(10):e0259012–e0259012.

80. Xie X, Xue Q, Zhou Y, Zhu K, Liu Q, Zhang J, et al. Mental Health Status Among Children in Home Confinement During the Coronavirus Disease 2019 Outbreak in Hubei Province, China. JAMA Pediatr. 2020;174(9):898–900.

81. Wang C, López-Núñez MI, Pan R, Wan X, Tan Y, Xu L, et al. The Impact of the COVID-19 Pandemic on Physical and Mental Health in China and Spain: Cross-sectional Study. JMIR Form Res. 2021;5(5):e27818–e27818.

82. Wang Q, Feng H, Wang M, Xie Y, Hou B, Lu X, et al. Mental Health and Psychological Responses During the Coronavirus Disease 2019 Epidemic: A Comparison Between Wuhan and Other Areas in China. Psychosom Med. 2021;83(4):322–7.

83. Xia Y, Kou L, Zhang G, Han C, Hu J, Wan F, et al. Investigation on sleep and mental health of patients with Parkinson’s disease during the Coronavirus disease 2019 pandemic. Sleep Med. 2020;75:428–33.

84. Wauters A, Vervoort T, Dhondt K, Soenens B, Vansteenkiste M, Morbée S, et al. Mental Health Outcomes Among Parents of Children With a Chronic Disease During the COVID-19 Pandemic: The Role of Parental Burn-Out. J Pediatr Psychol. 2022;47(4):420–31.

85. Wang H, Si MY, Su XY, Huang YM, Xiao WJ, Wang WJ, et al. [Mental Health Status and Its Influencing Factors among College Students during the Epidemic of Coronavirus Disease 2019:A Multi-center Cross-sectional Study]. Zhongguo Yi Xue Ke Xue Yuan Xue Bao. 2022;44(1):30–9.

86. Teng YM, Wu KS, Xu D. The Association Between Fear of Coronavirus Disease 2019, Mental Health, and Turnover Intention Among Quarantine Hotel Employees in China. Front Public Health. 2021;9:668774.

87. van Niekerk RL, van Gent MM. Mental health and well-being of university staff during the coronavirus disease 2019 levels 4 and 5 lockdown in an Eastern Cape university, South Africa. South Afr J Psychiatry. 2021;27:1589.

88. Vujčić I, Safiye T, Milikić B, Popović E, Dubljanin D, Dubljanin E, et al. Coronavirus Disease 2019 (COVID-19) Epidemic and Mental Health Status in the General Adult Population of Serbia: A Cross-Sectional Study. Int J Environ Res Public Health. 2021;18(4):1957.

89. Anjara SG, Bonetto C, Van Bortel T, Brayne C. Using the GHQ-12 to screen for mental health problems among primary care patients: psychometrics and practical considerations. Int J Ment Health Syst. 2020;14(1):62.

90. Boyles OM. How to Write Couples Therapy Notes [Internet]. 2022. Available from: https://www.icanotes.com/features/charting/assessment-tools/

91. McBride O, Murphy J, Shevlin M, Miller JG, Hartman TK, Hyland P, et al. An overview of the context, design and conduct of the first two waves of the COVID-19 Psychological Research Consortium (C19PRC) Study. 2020; Available from: https://psyarxiv.com/z3q5p/

92. Pmhealth NP. Screening Tools. https://pmhealthnp.com/screening-tools/. 2022.

93. Review HB. How to Measure Burnout Accurately and Ethically. https://hbr.org/2021/03/how-to-measure-burnout-accurately-and-ethically. 2021.

94. Jörns-Presentati A, Napp AK, Dessauvagie AS, Stein DJ, Jonker D, Breet E, et al. The prevalence of mental health problems in sub-Saharan adolescents: A systematic review. PLOS ONE. 2021;16(5):e0251689–e0251689.

95. Pieh C, Budimir S, Humer E, Probst T. Comparing Mental Health During the COVID-19 Lockdown and 6 Months After the Lockdown in Austria: A Longitudinal Study. Front Psychiatry [Internet]. 2021;12. Available from: https://pubmed.ncbi.nlm.nih.gov/33859579/

96. Pierce M, Hope H, Ford T, Hatch S, Hotopf M, John A, et al. Mental health before and during the COVID-19 pandemic: a longitudinal probability sample survey of the UK population. Lancet Psychiatry. 2020;7(10):883.

97. Robles R, Rodríguez E, Vega-Ramírez H, Álvarez-Icaza D, Madrigal E, Durand S, et al. Mental health problems among healthcare workers involved with the COVID-19 outbreak. Braz J Psychiatry. 2021;43(5):494–503.

98. Sampaio F, Sequeira C, Teixeira L. Nurses’ Mental Health During the Covid-19 Outbreak. J Occup Environ Med. 2020;62(10):783–7.

99. Serrano-Ripoll MJ, Meneses-Echavez JF, Ricci-Cabello I, Fraile-Navarro D, Fiol-deRoque MA, Pastor-Moreno G, et al. Impact of viral epidemic outbreaks on mental health of healthcare workers: a rapid systematic review and meta-analysis. J Affect Disord. 2020;277:347–57.

100. Albert PR. Why is depression more prevalent in women? J Psychiatry Neurosci JPN. 2015;40(4):219–21.

101. Goodman JH. Women’s mental health. JOGNN - J Obstet Gynecol Neonatal Nurs. 2005;34(2):245.

102. Mokhtari M, Dehghan SF, Asghari M, Ghasembaklo U, Mohamadyari G, Azadmanesh SA, et al. Epidemiology of mental health problems in female students: A questionnaire survey. J Epidemiol Glob Health. 2013;3(2):83.

103. Nzimande NP, El Tantawi M, Zuñiga RAA, Opoku-Sarkodie R, Brown B, Ezechi OC, et al. Sex differences in the experience of COVID-19 post-traumatic stress symptoms by adults in South Africa. BMC Psychiatry. 2022;22(1):1–8.

104. Serpytis P, Navickas P, Lukaviciute L, Navickas A, Aranauskas R, Serpytis R, et al. Gender-Based Differences in Anxiety and Depression Following Acute Myocardial Infarction. Arq Bras Cardiol. 2018;111(5):676–83.

105. Suanrueang P, Peltzer K, Suen MW, Lin HF, Er TK. Trends and Gender Differences in Mental Disorders in Hospitalized Patients in Thailand. Inq J Med Care Organ Provis Financ. 2022;59:1–14.

106. Oginni OA, Oloniniyi IO, Ibigbami O, Ugo V, Amiola A, Ogunbajo A, et al. Depressive and anxiety symptoms and COVID-19-related factors among men and women in Nigeria. PLOS ONE. 2021;16(8):e0256690–e0256690.

107. Demissie DB, Bitew ZW. Mental health effect of COVID-19 pandemic among women who are pregnant and/or lactating: A systematic review and meta-analysis. SAGE Open Med. 2021;9:205031212110261–205031212110261.

108. Chinvararak C, Kerdcharoen N, Pruttithavorn W, Polruamngern N, Asawaroekwisoot T, Munsukpol W, et al. Mental health among healthcare workers during COVID-19 pandemic in Thailand. PLOS ONE. 2022;17(5):e0268704–e0268704.

109. Ghaleb Y, Lami F, Al M, Hiba N, Rashak A, Samy S, et al. Mental health impacts of COVID-19 on healthcare workers in the Eastern Mediterranean Region: a multi-country study. J Public Health. 2021;43(Supplement_3):iii34.—iii42.

110. Blasco-Belled A, Tejada-Gallardo C, Fatsini-Prats M, Carles Alsinet ·. Mental health among the general population and healthcare workers during the COVID-19 pandemic: A meta-analysis of well-being and psychological distress prevalence. Curr Psychol 2022. 2022;1:1–12.

111. Baier AL, Marques L, Borba CPC, Kelly H, Clair-Hayes K, Dixon De Silva L, et al. Training needs among nonmental health professionals working with service members: A qualitative investigation. Mil Psychol Off J Div Mil Psychol Am Psychol Assoc. 2019;31(1):71.

112. Haverkamp FJC, van Leest TAJ, Muhrbeck M, Hoencamp R, Wladis A, Tan ECTH. Self-perceived preparedness and training needs of healthcare personnel on humanitarian mission: a pre- and post-deployment survey. World J Emerg Surg WJES. 2022;17(1):14.

113. Santomauro DF, Mantilla Herrera AM, Shadid J, Zheng P, Ashbaugh C, Pigott DM, et al. Global prevalence and burden of depressive and anxiety disorders in 204 countries and territories in 2020 due to the COVID-19 pandemic. The Lancet. 2021;398(10312):1700–12.

114. WHO. Mental Health and COVID-19: Early evidence of the pandemic’s impact [Internet]. World Health Organization Scientific brief. 2022. Available from: https://apps.who.int/iris/rest/bitstreams/1412184/retrieve

115. Hossain MM, Tasnim S, Sultana A, Faizah F, Mazumder H, Zou L, et al. Epidemiology of mental health problems in COVID-19: a review. F1000Research. 2020;9:636.

116. Alfaifi A, Darraj A, El-Setouhy M, Affairs JH. The Psychological Impact of Quarantine During the COVID-19 Pandemic on Quarantined Non-Healthcare Workers, Quarantined Healthcare Workers, and Medical Staff at the Quarantine Facility in Saudi Arabia. Psychol Res Behav Manag. 2022;15:1259–70.

117. Pancani L, Marinucci M, Aureli N, Riva P. Forced Social Isolation and Mental Health: A Study on 1,006 Italians Under COVID-19 Lockdown. Front Psychol. 2021;12:1540.

118. O’Caoimh R, O’Donovan MR, Monahan MP, Dalton O’Connor C, Buckley C, Kilty C, et al. Psychosocial Impact of COVID-19 Nursing Home Restrictions on Visitors of Residents With Cognitive Impairment: A Cross-Sectional Study as Part of the Engaging Remotely in Care (ERiC) Project. Front Psychiatry. 2020;11:1115.

119. Lawson K. How Do Thoughts and Emotions Affect Health? [Internet]. Earl E. Bakken Center for Spirituality and Healing. 2022. Available from: https://www.takingcharge.csh.umn.edu/how-do-thoughts-and-emotions-affect-health

120. Egelund T, Lausten M. Prevalence of mental health problems among children placed in out-of-home care in Denmark. Child Fam Soc Work. 2009;14(2):156–65.

121. Røsand GMB, Slinning K, Eberhard-Gran M, Røysamb E, Tambs K. The buffering effect of relationship satisfaction on emotional distress in couples. BMC Public Health [Internet]. 2012;12(1). Available from: https://pubmed.ncbi.nlm.nih.gov/22264243/

122. Holt-Lunstad J, Smith TB, Layton JB. Social Relationships and Mortality Risk: A Meta-analytic Review. PLOS Med. 2010;7(7):e1000316–e1000316.

123. Loades ME, Chatburn E, Higson-Sweeney N, Reynolds S, Shafran R, Brigden A, et al. Rapid Systematic Review: The Impact of Social Isolation and Loneliness on the Mental Health of Children and Adolescents in the Context of COVID-19. J Am Acad Child Adolesc Psychiatry. 2020;59(11):1218.

124. Novetney A. The risks of social isolation [Internet]. American Psychology Association, Monitor of Psychology. 2019. Available from: https://www.apa.org/monitor/2019/05/ce-corner-isolation

125. Simpson NJ, Oliffe JL, Rice SM, Kealy D, Seidler ZE, Ogrodniczuk JS. Social Disconnection and Psychological Distress in Canadian Men During the COVID-19 Pandemic. Am J Mens Health. 2022;16(1):155798832210781–155798832210781.

126. PsychGuides. Love and Mental Illness: A Survey of Psychological Well-Being and Intimate Partnerships [Internet]. PsychGuides. 2022. Available from: https://www.psychguides.com/interact/love-and-mental-illness/

127. Robb-Dover K. Red Flags That a Relationship Is Bad for Your Mental Health [Internet]. FHE Health Expert Columns. 2022. Available from: https://fherehab.com/learning/relationship-red-flag-mental-health

128. Falconier MK, Nussbeck F, Bodenmann G, Schneider H, Bradbury T. Stress from daily hassles in couples: its effects on intradyadic stress, relationship satisfaction, and physical and psychological well-being. J Marital Fam Ther. 2015;41(2):221–35.

129. Akbar Z, Aisyawati MS. Coping Strategy, Social Support, and Psychological Distress Among University Students in Jakarta, Indonesia During the COVID-19 Pandemic. Front Psychol. 2021;12:3409.

130. El Garhy NM, Hegazy G, Shata M, Ibrahim M, Kariem HA, Eleleedy A, et al. The Association between Emotional Intelligence and Depression among Medical Students in Suez Canal University. medRxiv. 2022;2022.08.31.22279446-2022.08.31.22279446.

131. Hacimusalar Y, Kahve AC, Yasar AB, Aydin MS. Anxiety and hopelessness levels in COVID-19 pandemic: A comparative study of healthcare professionals and other community sample in Turkey. J Psychiatr Res. 2020;129:181.

132. Harandi TF, Taghinasab MM, Nayeri TD. The correlation of social support with mental health: A meta-analysis. Electron Physician. 2017;9(9):5212.

133. Quinto RM, De Vincenzo F, Graceffa D, Bonifati C, Innamorati M, Iani L. The Relationship between Alexithymia and Mental Health Is Fully Mediated by Anxiety and Depression in Patients with Psoriasis. Int J Environ Res Public Health. 2022;19(6):3649.

134. Khalil M, Ghayas S, Adil A, Niazi S. Self-efficacy and Mental health among university students: Mediating role of assertiveness -. Rawal Med J. 2021;46(2):416–9.

135. Tahmassian K, Moghadam NJ. Relationship Between Self-Efficacy and Symptoms of Anxiety, Depression, Worry and Social Avoidance in a Normal Sample of Students. Iran J Psychiatry Behav Sci. 2011;5(2):91.

136. Hu T, Zhang D, Wang J, Mistry R, Ran G, Wang X. Relation between emotion regulation and mental health: a meta-analysis review. Psychol Rep. 2014;114(2):341–62.

137. Golberstein E, Professor A. The Effects of Income on Mental Health: Evidence from the Social Security Notch. J Ment Health Policy Econ. 2015;18(1):27.

138. Sareen J, Afifi TO, McMillan KA, Asmundson GJG. Relationship Between Household Income and Mental Disorders: Findings From a Population-Based Longitudinal Study. Arch Gen Psychiatry. 2011;68(4):419–27.

139. Shields-Zeeman L, Smit F. The impact of income on mental health. Lancet Public Health. 2022;7(6):e486.–e487.

140. CleavelandClinic. Chronic Illness and Depression: Causes, Symptoms, Treatment [Internet]. Cleaveland Clinic. 2021. Available from: https://my.clevelandclinic.org/health/articles/9288-chronic-illness-and-depression

141. Fernandez G. The Intersection of Mental Health and Chronic Disease [Internet]. John Hopkins Bloomberg School of Public Health. 2021. Available from: https://publichealth.jhu.edu/2021/the-intersection-of-mental-health-and-chronic-disease

142. Limone P, Toto GA. Factors That Predispose Undergraduates to Mental Issues: A Cumulative Literature Review for Future Research Perspectives. Front Public Health. 2022;10:831349.

143. Turner J, Kelly B. Emotional dimensions of chronic disease. West J Med. 2000;172(2):124.

144. MacMillan A. Why Mental Illness Can Fuel Physical Disease | Time [Internet]. Time Inc. 2017. Available from: https://time-com.cdn.ampproject.org/c/s/time.com/4679492/depression-anxiety-chronic-disease/?amp=true

145. MHF. Long-term physical conditions and mental health [Internet]. Mental Health Foundation. 2022. Available from: https://www.mentalhealth.org.uk/explore-mental-health/a-z-topics/long-term-physical-conditions-and-mental-health

146. Al-Aly Z. Mental health in people with covid-19. BMJ [Internet]. 2022;376. Available from: https://www.bmj.com/content/376/bmj.o415

147. Rapaport L. COVID-19 Patients at Increased Risk of Mental Health Issues [Internet]. Everyday Health. 2022. Available from: https://www.everydayhealth.com/coronavirus/covid-19-patients-at-increased-risk-for-mental-health-issues/

148. Salari N, Hosseinian-Far A, Jalali R, Vaisi-Raygani A, Rasoulpoor S, Mohammadi M, et al. Prevalence of stress, anxiety, depression among the general population during the COVID-19 pandemic: A systematic review and meta-analysis. Glob Health. 2020;16(1):1–11.

149. UNR. Impact of Social Media on Youth Mental Health [Internet]. University of Nevada, Reno. 2022. Available from: https://onlinedegrees.unr.edu/online-master-of-public-health/impact-of-social-media-on-youth-mental-health/

150. Zhao N, Zhou G. Social Media Use and Mental Health during the COVID-19 Pandemic: Moderator Role of Disaster Stressor and Mediator Role of Negative Affect. Appl Psychol Health Well-Being. 2020;12(4):1019.

151. Naslund JA, Bondre A, Torous J, Aschbrenner KA. Social Media and Mental Health: Benefits, Risks, and Opportunities for Research and Practice. J Technol Behav Sci 2020 53. 2020;5(3):245–57.

152. Sibonney C. Social Media Can Destroy Our Mental Health. What Can We Do About It? | SELF [Internet]. SELF. 2022. Available from: https://www-self-com.cdn.ampproject.org/c/s/www.self.com/story/social-media-mental-health-effects/amp

153. De Kock JH, Latham HA, Leslie SJ, Grindle M, Munoz SA, Ellis L, et al. A rapid review of the impact of COVID-19 on the mental health of healthcare workers: implications for supporting psychological well-being. BMC Public Health. 2021;21(1):1–18.

154. Baiyewu O, Elugbadebo O, Oshodi Y. Burden of COVID-19 on mental health of older adults in a fragile healthcare system: the case of Nigeria: dealing with inequalities and inadequacies. Int Psychogeriatr. 2020;32(10):1.

155. Bik-Multanowska K, Mikocka-Walus A, Fernando J, Westrupp E. Mental distress of parents with chronic diseases during the COVID-19 pandemic in Australia: A prospective cohort study. J Psychosom Res. 2022;152:110688.

156. Fekadu G, Bekele F, Tolossa T, Fetensa G, Turi E, Getachew M, et al. Impact of COVID-19 pandemic on chronic diseases care follow-up and current perspectives in low resource settings: a narrative review. Int J Physiol Pathophysiol Pharmacol. 2021;13(3):86.

157. Kompaniyets L, Pennington AF, Goodman AB, Rosenblum HG, Belay B, Ko JY, et al. Underlying Medical Conditions and Severe Illness Among 540,667 Adults Hospitalized With COVID-19, March 2020–March 2021. Prev Chronic Dis. 2021;18:1–13.

158. Shah K, Mann S, Singh R, Bangar R, Kulkarni R. Impact of COVID-19 on the Mental Health of Children and Adolescents. Cureus. 2020;12(8):e10051–e10051.

159. Wang Y, Shi L, Que J, Lu Q, Liu L, Lu Z, et al. The impact of quarantine on mental health status among general population in China during the COVID-19 pandemic. Mol Psychiatry 2021 269. 2021;26(9):4813–22.

160. OECD. Tackling the mental health impact of the COVID-19 crisis: An integrated, whole-of-society response [Internet]. OECD Policy Responses to Coronavirus. 2021. Available from: https://www.oecd.org/coronavirus/policy-responses/tackling-the-mental-health-impact-of-the-covid-19-crisis-an-integrated-whole-of-society-response-0ccafa0b/

161. OECD. Tourism Policy Responses to the coronavirus (COVID-19) [Internet]. OECD Policy Responses to Coronavirus. 2020. Available from: https://www.oecd.org/coronavirus/policy-responses/tourism-policy-responses-to-the-coronavirus-covid-19-6466aa20/

162. Schneidman W, Mkhize M. African governments ease COVID-19 restrictions and reopen economies | Cov Africa [Internet]. Cov Africa Current Affairs. 2020. Available from: https://www.covafrica.com/2020/10/african-governments-ease-covid-19-restrictions-and-reopen-economies/

